# Effect of Vaccination on Household Transmission of SARS-CoV-2 Delta VOC

**DOI:** 10.1101/2022.01.06.22268841

**Authors:** Frederik Plesner Lyngse, Kåre Mølbak, Matt Denwood, Lasse Engbo Christiansen, Camilla Holten Møller, Morten Rasmussen, Arieh Sierrah Cohen, Marc Stegger, Jannik Fonager, Raphael Sieber, Kirsten Ellegaard, Claus Nielsen, Carsten Thure Kirkeby

## Abstract

The SARS-CoV-2 Delta variant of concern (VOC), which has shown increased transmission compared with previous variants, emerged rapidly globally during the first half of 2021, and became one of the most widespread SARS-CoV-2 variants worldwide. We utilized total population data from 24,693 Danish households with 53,584 potential secondary cases to estimate household transmission of the Delta VOC in relation to vaccination status. We found that the vaccine effectiveness against susceptibility (VES) was 61% (95%-CI: 59-63) and that the vaccine effectiveness against transmissibility (VET) was 42% (95%-CI: 39-45). We also found that unvaccinated individuals with an infection exhibited a higher viral load (one third of a standard deviation) compared to fully vaccinated individuals with a breakthrough infection. Our results imply that vaccinations reduce susceptibility as well as transmissibility. The results are important for policy makers to select strategies for reducing transmission of SARS-CoV-2.

## 2 Introduction

The current SARS-CoV-2 pandemic is of major concern worldwide, and vaccination for SARS-CoV-2 is a central part of the strategy to control the epidemic. Nevertheless, pandemic control is being challenged by the ability of SARS-CoV-2 to continuously evolve into new genomic variants of concern (VOC) with differing characteristics in terms of transmissibility and immune evasion. During 2021, the Delta VOC (SARS-CoV-2 Lineage B.1.617.2) became a major concern, due to the evidence for higher transmissibility compared to previous variants (Liu & Rocklöv, 2021; Campbell et al., 2021; Lopez Bernal et al., 2021; Dagpunar, 2021; Allen et al., 2021). Furthermore, some studies suggest that the Delta VOC possess increased immune evasion properties relative to previous variants, causing a higher number of breakthrough infections in vaccinated individuals (Farinholt et al., 2021; Mlcochova et al., 2021). During the second half of 2021, the Delta VOC caused new surges in COVID-19 infections and hospitalizations in several countries, putting a pressure on health care systems despite broad vaccination rollout. In response, several countries reintroduced restrictions and non-pharmaceutical interventions (NPIs) to sustain epidemic control. To understand the full potential of the Delta VOC, it is essential to estimate the vaccine effectiveness for this variant. It is inherently difficult to estimate real-world vaccine effectiveness (VE), as vaccination roll-out has prioritized special risk-groups, and take-up has been large, causing potential bias in the control group of unvaccinated individuals. Furthermore, waning immunity complicates the identification of the VE.

It is therefore essential to quantify the vaccine effectiveness for susceptibility (VES), the vaccine effectiveness for transmissibility (VET), and the combined effect (VEC). Harris et al. (2021) studied the effect of first vaccination and found that transmission from vaccinated individuals was 40% to 50% lower than from unvaccinated individuals. They pointed out that more data and studies are needed to estimate the reduction in transmission for fully vaccinated individuals.

There are several issues that make it difficult to quantify the VES and VET. In principle, an infection with SARS-CoV-2 generally requires three things: i) an infected primary case that is able to transmit the virus, ii) a potential secondary case that is susceptible to the virus, and iii) a contact/transmission event between the primary and the potential secondary case. Vaccinations affect the susceptibility of the potential secondary case, but may also affect the transmissibility of a primary case, conditional on a breakthrough infection overcoming the vaccinated status of the individual. Furthermore, individuals may change their behavior due to the vaccination status of themselves and/or others, complicating the process of separating the effects of VES and VET.

For policy makers aiming to control the pandemic, vaccinations play a key role. It is essential to quantify the vaccine effectiveness—–not just against susceptibility—but also against transmissibility.

In this study we used national population data to estimate vaccine effectiveness within Danish households. We estimate two key parameters: Vaccine effectiveness against susceptibility (VES) and vaccine effectiveness against transmissibility (VET) for the Delta VOC to and from individuals with and without vaccination against SARS-CoV-2.

## 3 Data and Methods

### 3.1 Study period

We selected the study period to include primary cases with the Delta VOC, which became dominant in Denmark around the middle of July 2021. We used data where the primary case within each household was between 21 June 2021 to 26 October 2021. Potential secondary cases were followed up to 9 November to provide sufficient time for them to subsequently test positive. Appendix 6 provides background information on the study period, including the number of tests performed, the number of cases, and vaccination roll-out.

### 3.2 Data

This study was conducted using Danish register data. In Denmark, all citizens have a personal identification number that allows their information to be linked across different data registers at the individual level. We linked all members of the same household via their registered home address and merged this with data on all RT-PCR tests for SARS-CoV-2 from the Danish Microbiology Database (MiBa). We identified the first positive test result in each household and defined the corresponding individual as the primary case. All other household members were defined as potential secondary cases. Information on vaccinations of each individual was obtained from the Danish Vaccination Register (DDV) as described below. We only considered households with 2-6 members in order to exclude e.g. student dorms, social housing, and care facilities. We only included households where the primary case tested positive with the Delta VOC and where no other household member tested positive on the same day. Secondary cases were defined as all cases testing positive within 1-14 days of the primary case within each household.

Individuals were classified depending on their vaccination status on the test-positive day of the primary case. Individuals who had not received a first dose were classified as not vaccinated. The definitions of full vaccinations were: Comirnaty (Pfizer/BioNTech): 7 days after second dose; Vaxzevria (AstraZeneca): 15 days after second dose; Spikevax (Moderna): 14 days after second dose; COVID-19 VACCINE Janssen (Johnson & Johnson): 14 days after vaccination. If an individual was cross vaccinated (mainly first dose of Vaxzevria and second dose of Comirnaty), the definition of the second dose vaccination was used. Individuals that were in the period between the first dose and fully vaccinated were defined as partially vaccinated and excluded. Any individual that had received a booster vaccination was also excluded. Lastly, all households with a previous infection (positive RT-PCR test) were excluded.

### 3.3 Statistical analyses

We defined the overall household secondary attack rate (SAR) as the proportion of potential secondary cases that tested positive between 1-14 days following the identification of the primary case within the same household.

We defined the combined vaccine effectiveness (VEC, the combined effect of vaccination on both the infected primary case and the potential secondary case) as one minus the relative risk (RR) of vaccinated individuals compared to unvaccinated individuals, following Halloran et al. (2003).

To estimate the vaccine effectiveness, we used a generalized linear model (GLM), with Poisson distribution response and a log link function, comparing fully vaccinated individuals with unvaccinated individuals (see below). The modified GLM routine also included standard errors clustered on the household level. The use of a Poisson distribution to describe a binary response was to facilitate estimation of relative risks rather than odds ratios. The regression model included fixed effects controls for age (categorical effects in 5-year age groups) and sex of both the primary and potential secondary cases, and fixed effects for household size (categorical effects). We also included calendar week fixed effects to control for temporal variation, e.g., behavior, changes in restrictions, vaccination coverage and overall incidence.

To estimate the extent to which vaccination reduces susceptibility to infection (VES), we estimated the relative risk (RR) of the SAR for potential secondary cases that were fully vaccinated compared to the SAR for the potential secondary cases that were not vaccinated. To separate the effect of vaccination affecting susceptibility (VES) from transmissibility (VET), we also stratified by vaccination status of the primary case.

To estimate the extent to which vaccination protects against transmissibility to other household members, i.e., the VET, we estimated the RR of the SAR for primary cases that were fully vaccinated compared to the SAR for primary cases that were not vaccinated. To separate the effect of vaccination affecting transmissibility (VET) from susceptibility (VES), we also stratified by vaccination status of the potential secondary case.

To estimate the combined effect of vaccine protection against susceptibility and transmissibility, we estimated the RR of the SAR for primary cases and potential secondary cases that were fully vaccinated compared to primary cases and potential secondary cases that were not vaccinated.

To explore the effect of vaccinations on infectiousness, we investigated the difference in viral load (proxied by Ct values) for vaccinated and unvaccinated secondary cases testing positive on the same day after exposure.

A more detailed description of the statistical methods is provided in Appendix 8.

#### 3.3.1 Additional analyses

To test the robustness of our main results, we conducted a number of additional analyses. To investigate the age related transmission patterns, we estimated the SAR, VET, VES and VEC stratified by the age of primary case and potential secondary cases. To investigate if secondary cases were due to household or community transmission, we investigated the intra-household correlation of subtype lineages of the Delta variant. We investigated the intra-household correlation of vaccination status, as the vaccination status between household members are likely correlated. We also investigated the sample Ct values from unvaccinated and fully vaccinated primary cases. Because a positive test result is provided conditionally on actually having a test, we investigated the probability of being tested across vaccination status of both the primary and potential secondary case. To investigate the robustness of our main results, we estimated the VES, VET and VEC, conditional on the potential secondary case having a test result. To investigate the effect of waning immunity, we estimated the VES, VET and VEC by time intervals since vaccination. Finally, to investigate if the VES, VET and VEC estimates are driven by a change in the viral load of the primary case, we estimated these while adjusting for the viral load of the primary case, using the sample Ct value as a proxy.

The results of the additional analyses are presented in Appendix 7.

### 3.4 Ethical statement

This study was conducted on administrative register data. According to Danish law, ethics approval is not needed for this type of research. All data management and analyses were carried out on the Danish Health Data Authority’s restricted research servers with project number FSEID-00004942. The publication only contains aggregated results and no personal data. The publication is, therefore, not covered by the European General Data Protection Regulation.

### 3.5 Data availability

The data used in this study are available under restricted access due to Danish data protection legislation. The data are available for research upon reasonable request to The Danish Health Data Authority and Statens Serum Institut and within the framework of the Danish data protection legislation and any required permission from Authorities. We performed no data collection and performed no sequencing specifically for this study.

## 4 Results

Table 1 shows the summary statistics of the data included in this study. The study included 24,693 primary cases, of which 33% (8,262) were fully vaccinated and 67% (16,431) were not vaccinated, and 53,584 potential secondary cases, of which 49% (26,098) were fully vaccinated and 52% (27,486) were not vaccinated. The SAR was generally lower among vaccinated potential secondary cases than among unvaccinated (Table 1). Lastly, for children between 0-10 years of age, we found a SAR at 27%.

**Table 1:** Summary Statistics

|  | Fully vaccinated |  |  |  | Unvaccinated |  |  |  |
| --- | --- | --- | --- | --- | --- | --- | --- | --- |
|  | Primary | Potential | Positive | Secondary | Primary | Potential | Positive | Secondary |
|  | Cases | Secondary | Secondary | Attack | Cases | Secondary | Secondary | Attack |
|  | Cases | Cases | Cases | Rate (%) | Cases | Cases | Cases | Rate (%) |
| **Total** | 8,262 | 26,098 | 3,816 | 15 | 16,431 | 27,486 | 7,815 | 28 |
| **Sex** |  |  |  |  |  |  |  |  |
| Male | 4,001 | 12,868 | 1,711 | 13 | 8,301 | 13,801 | 3,626 | 26 |
| Female | 4,261 | 13,230 | 2,105 | 16 | 8,130 | 13,685 | 4,189 | 31 |
| **Age** |  |  |  |  |  |  |  |  |
| 0-10 | 0 | 0 | 0 |  | 3,668 | 11,687 | 3,198 | 27 |
| 10-20 | 721 | 3,482 | 181 | 5 | 4,859 | 5,742 | 1,720 | 30 |
| 20-30 | 1,368 | 2,633 | 224 | 9 | 4,131 | 3,756 | 983 | 26 |
| 30-40 | 1,131 | 4,434 | 730 | 16 | 2,316 | 3,014 | 1,028 | 34 |
| 40-50 | 1,676 | 7,961 | 1,210 | 15 | 923 | 2,015 | 585 | 29 |
| 50-60 | 1,601 | 4,808 | 686 | 14 | 407 | 994 | 247 | 25 |
| 60-70 | 1,085 | 1,893 | 445 | 24 | 109 | 211 | 42 | 20 |
| 70-80 | 680 | 887 | 340 | 38 | 18 | 67 | 12 | 18 |
| **Household** |  |  |  |  |  |  |  |  |
| **Size** |  |  |  |  |  |  |  |  |
| 2 | 3,859 | 4,832 | 1,209 | 25 | 3,704 | 2,731 | 812 | 30 |
| 3 | 1,809 | 5,660 | 702 | 12 | 4,092 | 5,132 | 1,402 | 27 |
| 4 | 1,785 | 9,246 | 1,204 | 13 | 5,080 | 9,416 | 2,801 | 30 |
| 5 | 643 | 4,980 | 572 | 11 | 2,762 | 7,260 | 2,029 | 28 |
| 6 | 166 | 1,380 | 129 | 9 | 793 | 2,947 | 771 | 26 |
| **Vaccination** |  |  |  |  |  |  |  |  |
| Vaxzevria | 514 | 1,398 | 229 | 16 |  |  |  |  |
| Janssen | 529 | 546 | 108 | 20 |  |  |  |  |
| Spikevax | 366 | 2,441 | 243 | 10 |  |  |  |  |
| Cormirnaty | 6,853 | 21,713 | 3,236 | 15 |  |  |  |  |

Notes: The secondary attack rate (SAR) is expressed in percentages. Primary cases and potential secondary cases are here shown by groups of sex, age, household size and vaccination status, independent of each other. Appendix Table 3 provides summary statistics for potential and positive secondary cases are grouped based on the primary case characteristics.

Table 2 shows the vaccine effectiveness (VE) estimates. The pooled vaccine effectiveness against susceptibility (VES) was 61% (95%-CI: 59-63). The VES was 61% (95%-CI: 59-63) when the primary case was unvaccinated, compared to 46% (95%-CI: 40-52) when the primary case was fully vaccinated primary cases. The pooled vaccine effectiveness against transmissibility (VET) was 42% (95%-CI: 39-45). The VET was 31% (95%-CI: 26-36) when the potential secondary was for unvaccinated, compared to 10% (95%-CI: 0-18) when the potential secondary case was fully vaccinated. The combined effect (VEC) was 66% (95%-CI: 63-68), i.e., when both the primary case and potential secondary case were fully vaccinated compared to them being unvaccinated. Note that the VE estimates across columns are not directly comparable as they are estimated on stratified samples.

**Table 2:**
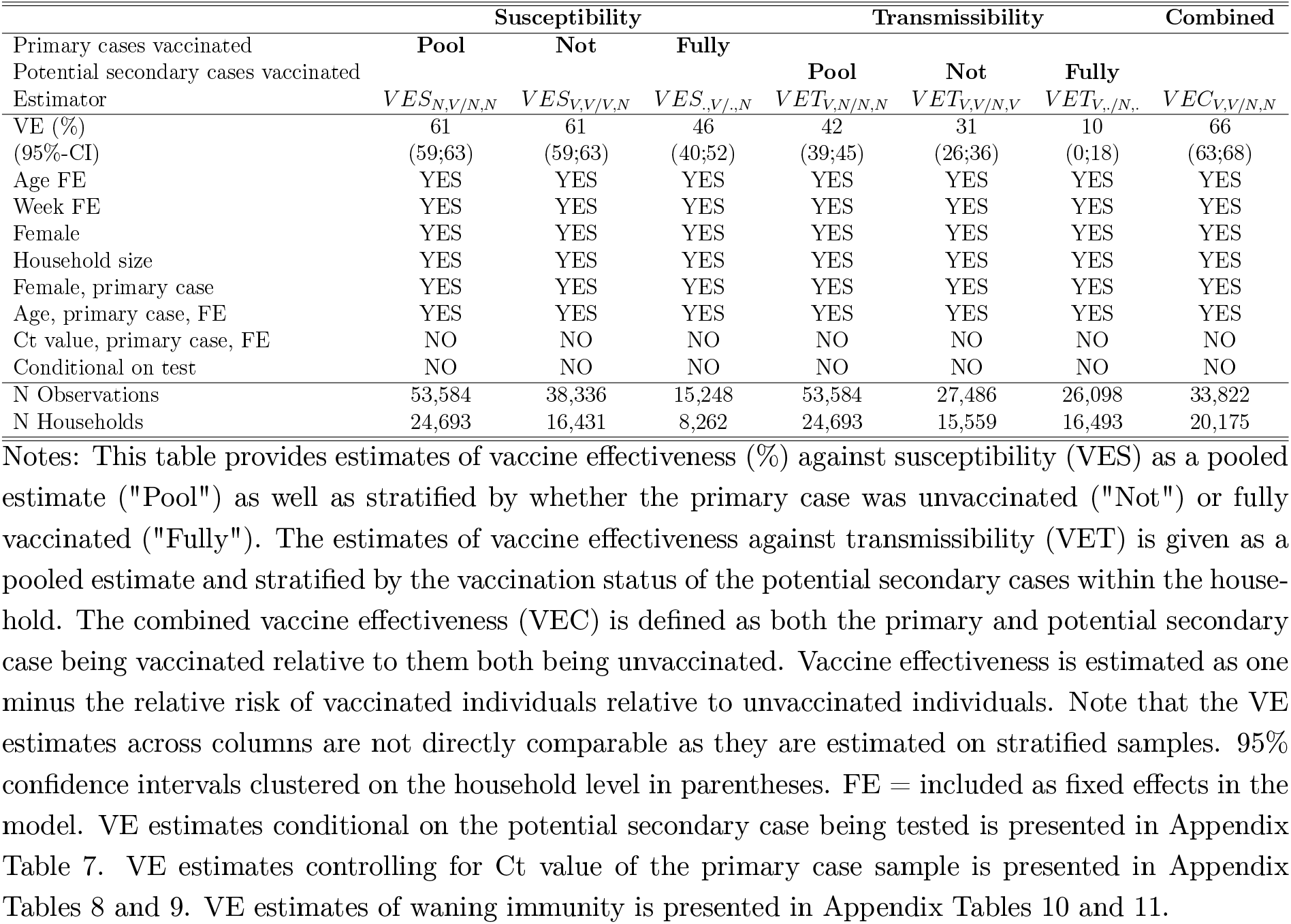
Vaccine effectiveness (%)

**Table 3:**
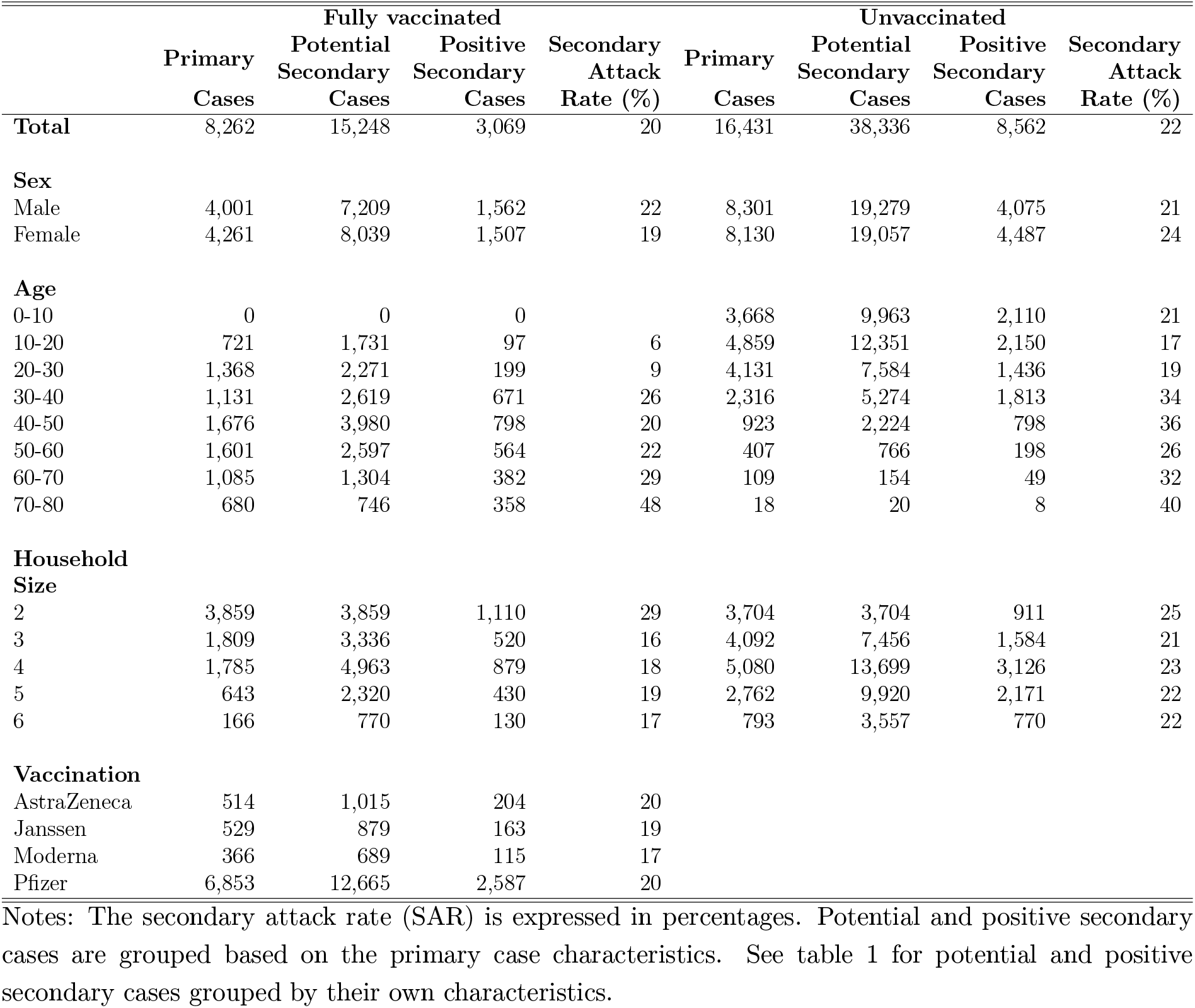
Summary Statistics, stratified by primary case level

Further robustness of the VE estimates are presented in Appendix 7.6. Tables 10 and 11 show estimates of VE since time of vaccination, i.e., waning immunity. The VES and VET decreased from 71% (95%-CI: 69-72) and 57% (95%-CI: 53-61), respectively, to 32% (95%-CI: 16-45) and 29% (95%-CI: 14-41), respectively, between time points corresponding to 0-1 months and 7-8 months after vaccination (Table 10).

Vaccinated secondary cases have a significantly lower viral load (higher Ct value) compared to unvaccinated secondary cases, independent of the day of testing after identification of the primary case (Figure 1). Vaccinated secondary cases have an increased Ct value of 1.6, corresponding to a third of a standard deviation (Appendix Table 6).

**Figure 1:**
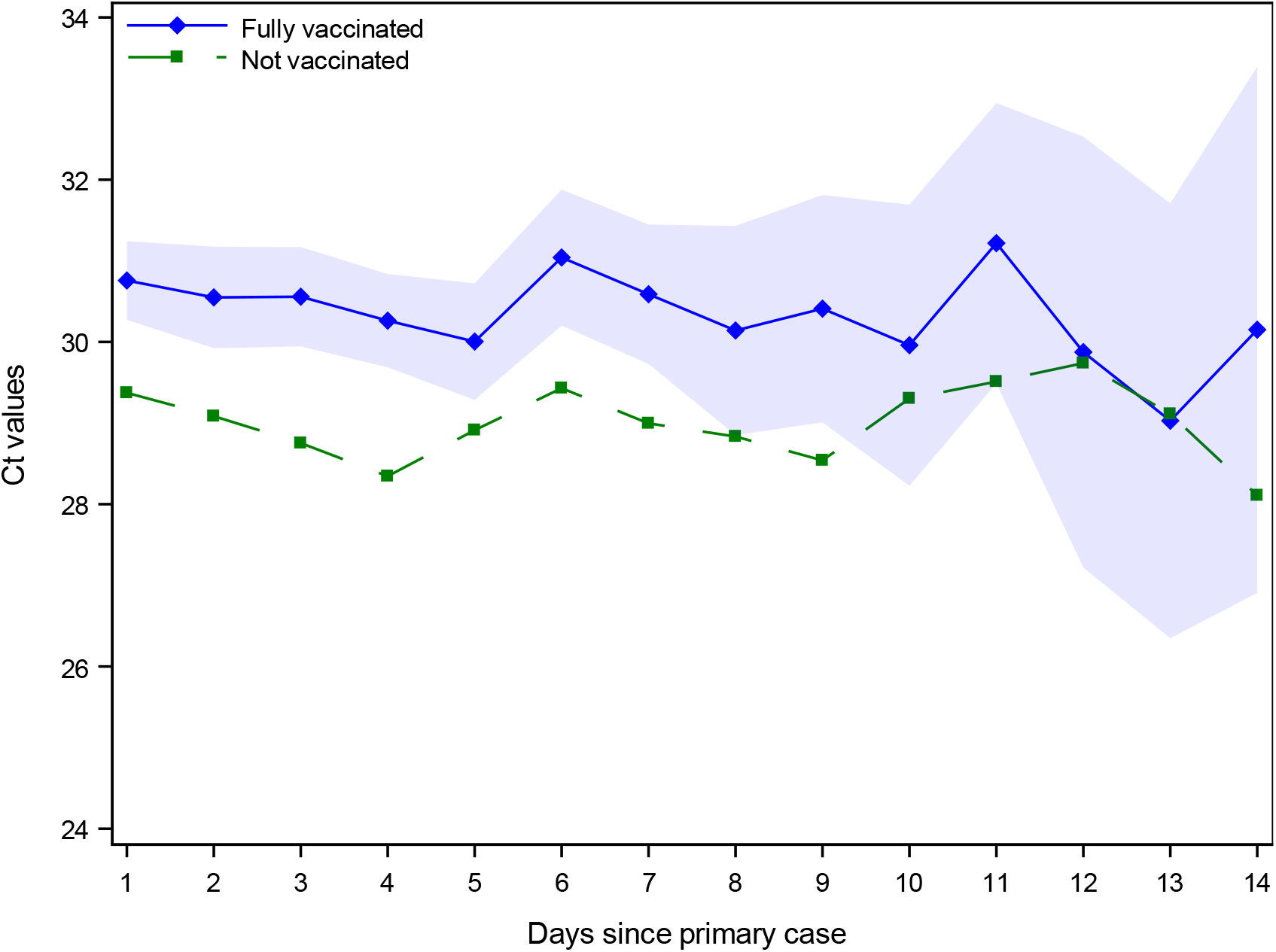
Ct values across vaccinated and unvaccinated positive secondary cases Notes: The Ct values in samples for positive secondary cases of unvaccinated and fully vaccinated follow the same pattern, indicating that the higher Ct values for fully vaccinated individuals is consistent and unrelated to the time of testing positive. Regression estimates include age fixed effects. Shaded areas are 95%-confidence intervals clustered on the household level. Appendix Table 6 provides regression estimates of the increased Ct values for vaccinated positive secondary cases.

## 5 Discussion and Conclusion

We used Danish national population data to estimate household transmission of SARS-CoV-2 Delta VOC to and from unvaccinated and fully vaccinated individuals. We found a vaccine effectiveness against susceptibility (VES) against the Delta VOC of 61% (95%-CI: 59-63), when the primary case was not vaccinated, and a VES of 46% (95%-CI: 40-52), when they were fully vaccinated. Furthermore, we found a vaccine effectiveness against transmissibility (VET) of 31% (95%-CI: 26-36), when the potential secondary case was not vaccinated, and 10% (95%-CI: 0-18), when they were fully vaccinated. Lastly, we found a combined vaccine effectiveness (VEC) of 66% (95%-CI: 63-68), when both the primary and potential secondary case were fully vaccinated.

Other studies used contact tracing data and found similar estimates. In the Netherlands, de Gier et al. (2021) found a household VET of 63% (95%-CI: 46–75) among unvaccinated potential secondary cases and a VET of 40% (95%-CI: 20–54) among fully vaccinated potential secondary cases. In Singapore, Ng et al. (2021) found a pooled VES of 62% (95%-CI: 38-80). Moreover, in a recent study, using the same approach as the present study, Lyngse et al. (2021b) showed a general higher VET for unvaccinated cases and a lower VET for booster-vaccinated cases, compared to fully vaccinated cases. They also found no significant difference in the VET for cases infected with the Omicron VOC compared to the Delta VOC.

It has previously been shown that vaccination protects against infection and hospitalization duration (Veneti et al., 2021; Levine-Tiefenbrun et al., 2021), severity of disease (Ruian Ke, 2021), and mortality (Mor et al., 2021) caused by the Delta VOC (Chia et al., 2021). We have demonstrated that vaccinations further protect against transmission, conditional on having a breakthrough infection. This protective effect is important, as some studies have indicated that the Delta VOC has increased immune evasion properties relative to other variants, causing a higher number of breakthrough infections in vaccinated individuals (Farinholt et al., 2021). This ability may be caused by a higher replication within the airway cells (Mlcochova et al., 2021).

Lower viral loads in vaccinated cases compared to unvaccinated cases could be a potential mechanism for this reduction in transmissibility. However, there is currently no agreement on the effect of vaccination on the viral load. Some studies found that vaccinations did not reduce the viral load of cases with a breakthrough infection. Chia et al. (2021), for example, found that vaccinated and unvaccinated individuals infected with the Delta VOC had similar Ct values at diagnosis, but that the viral loads decreased faster in vaccinated individuals. Contrary to this, we found that vaccinated cases had lower viral loads (higher Ct values), corroborating the findings of Levine-Tiefenbrun et al. (2021), who found that breakthrough infections with the Delta VOC resulted in lower viral loads in vaccinated cases compared with unvaccinated cases.

Our study found that the pooled VES generally decreased from 71% (95%-CI: 69-72) at 0-1 months after vaccination, compared to 32% (95%-CI: 16-45) 7-8 months after vaccination (Appendix Table 10). Similarly, the pooled VET generally decreased from 57% (95%-CI: 53-61) at 0-1 months after vaccination, compared to 29% (95%-CI: 14-41) 7-8 months after vaccination. This highlights the need for booster vaccinations in order to improve protection and reduce transmission.

Our results have important implications for policy makers. First, we have shown that vaccination reduces the transmission between infected and susceptible individuals, corroborating existing studies. Vaccination can therefore be used to reduce the risk of infection. Second, we found that vaccinations protect more against susceptibility than transmissibility. Therefor NPIs acting mostly on the transmissibility, such as wearing masks (Abaluck et al., 2021), can be necessary even for vaccinated individuals, especially when close to unvaccinated individuals. On the other hand, the reduced susceptibility of vaccinated individuals may facilitate the relaxation of guidelines among vaccinated individuals, e.g., at mass gatherings. This suggests that immunity passports can be an effective measure to reduce transmission, as previously suggested (Brown et al., 2020). Third, we showed that vaccination also protects against transmissibility, conditional on having a breakthrough infection. This indicates that prioritizing the vaccination of groups that have many contacts or work with vulnerable individuals—e.g., nursing home staff—is important for pandemic control. Fourth, our estimates can be used to inform simulation models of the current pandemic, which crucially rely on the parameters of susceptibility and transmissibility. As vaccination affects both the susceptibility and transmissibility, accurate estimates of these effects are critical to the models that are used to inform decision makers.

We here found a substantial degree of transmission to and from children. Unvaccinated children aged 0-10 years were susceptible to the SARS-CoV-2 Delta VOC with a SAR of 27% (Table 1). Furthermore, we found that unvaccinated children aged 0-20 years that were primary cases were able to infect vaccinated household members aged 20-60 with a SAR of 15% (Figure 7).

This study has several strengths. First, we combined several national-level datasets in order to control for individual specific factors relating to both the primary and potential secondary case. This is unique because standard approaches to estimating VE usually focus on VES or a combined measure. Second, throughout the study period, Denmark had a large testing capacity: testing was free and widely used. Third, all positive cases were being contacted by the official contact tracing unit. All household members are per definition close contacts and recommended to become RT-PCR tested twice after the identification of the primary case—unconditional on the vaccination status. Forth, Denmark has a high vaccination uptake across all vaccination groups. Overall, this provides a setting that allows us to estimate vaccine effectiveness both from and to vaccinated and unvaccinated individuals. Lyngse et al. (2021a) used data from whole genome sequencing to validate the same methods as used in the present study. They found an intra-household correlation of lineages between primary and positive secondary cases of 96-99%, indicating that most secondary cases were infected with the same lineage as the primary case. In the present study, the Delta VOC comprised more than 95% of all cases in society (Figure 3), making it impossible to investigate variation across different variants. However, using the subtype lineages of the Delta VOC, we found no significant difference in the intra-household correlation of lineages across vaccinated and unvaccinated individuals (Appendix 7.2). The data were collected while the Danish vaccination program was rolled out to all above 12 years of age. Therefore, unvaccinated individuals mainly represents individuals who were not yet invited for vaccination. This is a major strength of our study because a self-selected unvaccinated group might introduce a bias due to the fact that reluctancy to receive immunizations may correlate with other types of behavior.

**Table 4:**
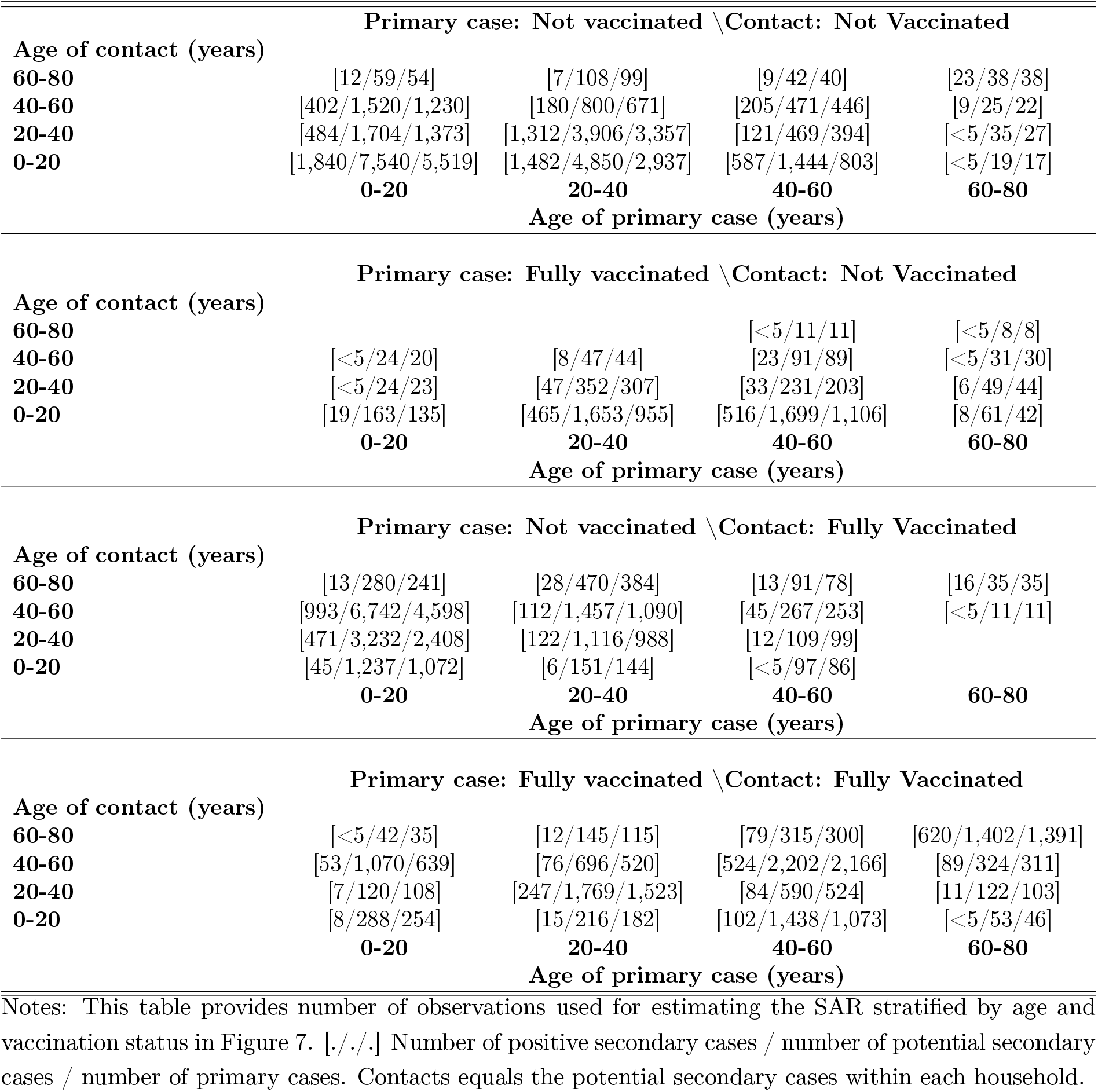
Number of observations for Figure 7

**Table 5:** Intra-household correlation of lineages

|  |  | Secondary Case |  |
| --- | --- | --- | --- |
|  | Pool | Vaccinated | Unvaccinated |
| Primary Case |  |  |  |
| Pool | 88<br>(87-89)<br>[9,580/6,852] | 89<br>(88-90)<br>[3,156/2,860] | 88<br>(87-89)<br>[6,424/4,652] |
| Vaccinated | 90<br>(88-91)<br>[2,443/1,980] | 89<br>(87-91)<br>[1,573/1,499] | 90<br>(88-92)<br>[870/656] |
| Unvaccinated | 88<br>(87-89)<br>[7,137/4,872] | 89<br>(87-90)<br>[1,583/1,361] | 87<br>(86-88)<br>[5,554/3,996] |

Notes: This table provides estimates on the intra-household correlation of lineages, i.e., the probability that the positive secondary case had the same subtype lineage as the primary case. 95%-confidence intervals clustered on the household level presented in parenthesis. Number of observations and household clusters for each point estimate are presented in hard brackets: [Number of observations/number of households].

Some limitations apply to this study. Firstly, we did not have access to clinical information. Secondly, vaccination status may affect the behavior of both primary cases and potential secondary cases, as they may adhere less to non-pharmaceutical interventions (NPIs) once they or others are vaccinated. Thus, the estimates in our study reflect both the biological aspect of susceptibility and transmissibility, as well as the behavioral aspects, and does not confer any information regarding severity of illness, which is expected to be lower among vaccinated individuals.

There are several behavioral aspects relating to vaccination. Once a primary case is identified within the household, the other members might allocate their time with that individual, conditional on their own vaccination status. For example, if the primary case is a child and one parent is fully vaccinated, while the other is not, then the family may choose to allocate the majority of childcare during the infection to the vaccinated parent, as they have a lower risk of being infected. This changed contact pattern may lead to a higher rate of infections in vaccinated individuals in households where there are also unvaccinated members. Furthermore, vaccinated individuals may in general adhere less to non-pharmaceutical interventions, such as keeping distance, wearing masks, etc. due to a perceived lower susceptibility and transmissibility. Unvaccinated individuals may be identified earlier in their infection than vaccinated individuals, as they are being more regularly tested. However, in Denmark, all contacts are encouraged to be PCR tested— regardless of their vaccination status—when a positive individual is found within the same household. We tested the difference of the probability of being tested after exposure between fully vaccinated and unvaccinated potential secondary cases. We found that vaccinated potential secondary cases were about 7 percentage points (10%) more likely to be tested compared to unvaccinated individuals (Appendix 7.4). This suggests that there are differences across the two groups that we cannot fully control for, e.g., general compliance to NPIs. These results are in line with the results in Nielsen & Petersen (2021) that show that unvaccinated individuals are less likely to adhere to government recommendations. This also implies that our VE estimates are a lower bound. When we restrict our analyses to only include individuals with a test result, we obtain higher VE estimates (Appendix Tables 7, 9, and 11). Moreover, there is likely a correlation of vaccination status between household members. i) Individuals living together may be more likely to share the same belief, for instance towards vaccination. ii) Individuals are likely to have a partner around their own age. As vaccination roll-out is based on age, household members are likely to be eligible for vaccination around the same calendar time. iii) There may be a fixed cost of being vaccinated, e.g., the travel from the home to the vaccination location. Thus, a household might pool their day of vaccination together to minimize travel costs. We found an intra-household correlation of vaccination status of 0.72 for individuals above age 12 years (Appendix 7.3).

Estimating VES and VET include several challenges. When few individuals are vaccinated in a population, it is less complicated to estimate the approximate vaccine effectiveness against susceptibility (VES), because their exposure can be assumed to come from unvaccinated cases. As vaccinations are rolled out in a country, the proportion of the population that is vaccinated increases. As a consequence, the proportion of contacts with vaccinated individuals also increases. Therefore, the real-life observed VES estimates are a composition of exposure from and to both vaccinated and unvaccinated individuals. If vaccination not only protects against infection, but also against transmission, the estimates are a combined effect of both VES and VET. The VET becomes increasingly more important, as vaccination rates throughout society increase. Furthermore, it is necessary to link primary cases to exposed contacts in order to estimate the VET.

Age is correlated with susceptibility and transmissibility as well as eligibility and roll-out of vaccinations. This implies a natural imbalance in the number of primary and potential secondary cases across age groups. To address this, we provide estimates of SAR and VE in all combinations of age groups, stratified by vaccination status of both the primary and potential secondary case (Appendix 7.1).

The estimates of VES are probably conservative compared with the general VES at the overall population level. Transmission within household is associated with more intense exposure than in the community in general, and since there is a relation between the degree of exposure and likelihood of breakthrough infection, it is expected that vaccines may work better in the community than in households. Secondly, the estimates based on an infection in a fully vaccinated individual is naturally conditioned on a breakthrough infection. The virus variant that has caused this infection may have been adapted to vaccine derived immunity, and may therefore be more likely to result in another breakthrough infection.

In conclusion, we have demonstrated that vaccination was able to reduce the transmission of the Delta VOC in Danish households over our study period. We estimated a vaccine effectiveness against susceptibility (VES) of 61% (95%-CI: 59-63) among unvaccinated primary cases and a vaccine effectiveness against transmissibility (VET) of 31% (95%-CI: 26-36) among unvaccinated potential secondary cases. This implies that vaccinations reduce susceptibility as well as transmissibility.

## Aknowledgements

We thank Statens Serum Institut and The Danish Health Data Authority for data access. We also thank the rest of the Expert Group for Mathematical Modelling of COVID-19 at Statens Serum Institut for helpful discussions, as well as as the Danish Covid-19 Genome Consortium for typing of positive COVID-19 samples.

## Contributions

FPL performed data analysis and produced all estimates and figures presented. FPL, CTK and KM wrote the first draft. MD contributed to the selection and description of statistical methods. All other authors contributed to the discussion and writing the final draft. All authors read and approved the final version of the manuscript.

### Supplementary Appendix

#### 6 Background

This section provides some background statistics on the SARS-CoV-2 pandemic situation in Denmark, between June and November, 2021.

Figure 2 shows the number of tests performed per million Danish citizens and the number of new positive cases identified per thousand Danish citizens between June and November, 2021.

**Figure 2:**
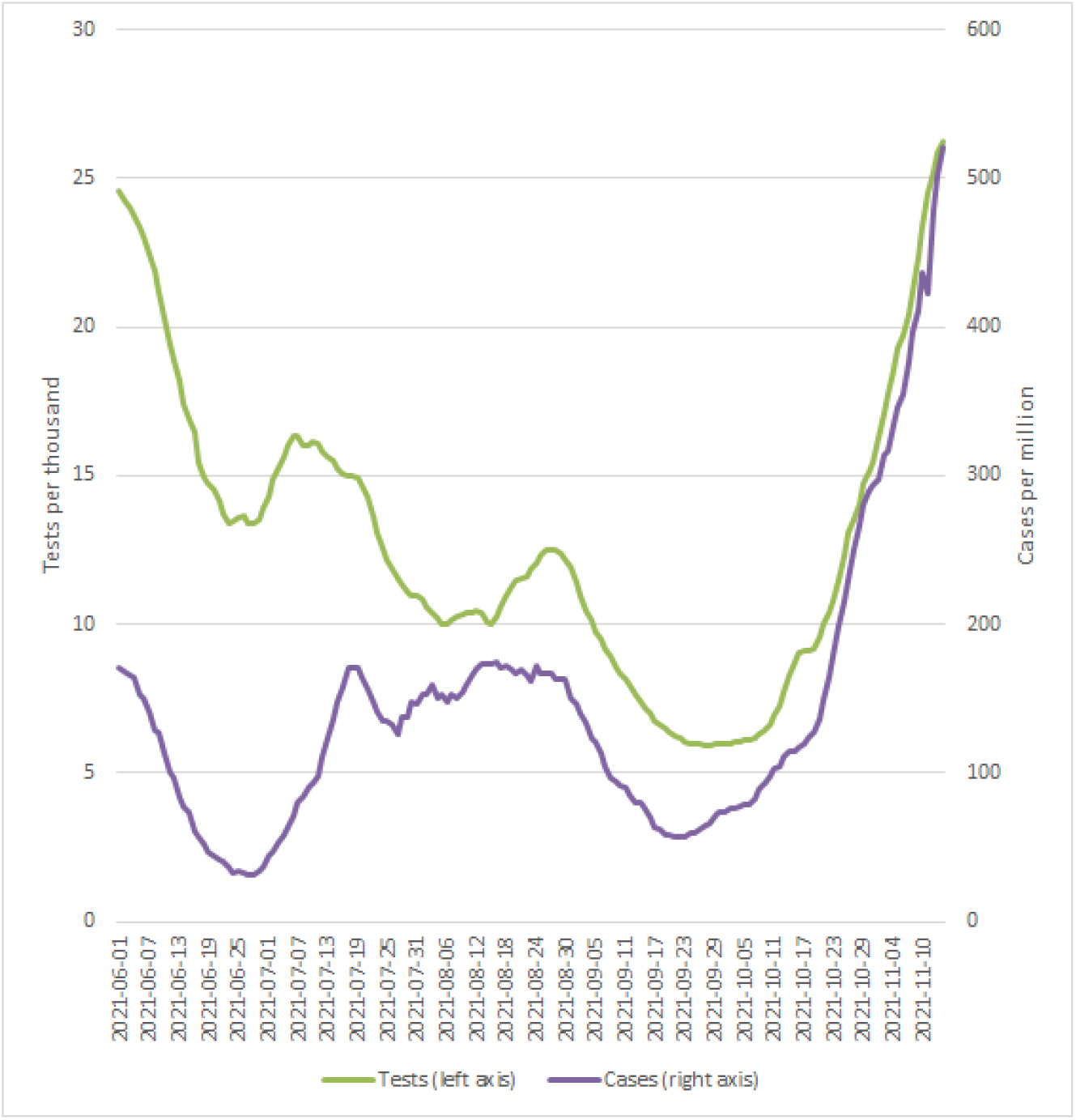
Tests and Cases Notes: This figure shows the number of tests performed per million Danish citizens and the number of new positive cases identified per thousand Danish citizens, June to November, 2021. Data source: SSI (2021a).

**Figure 3:**
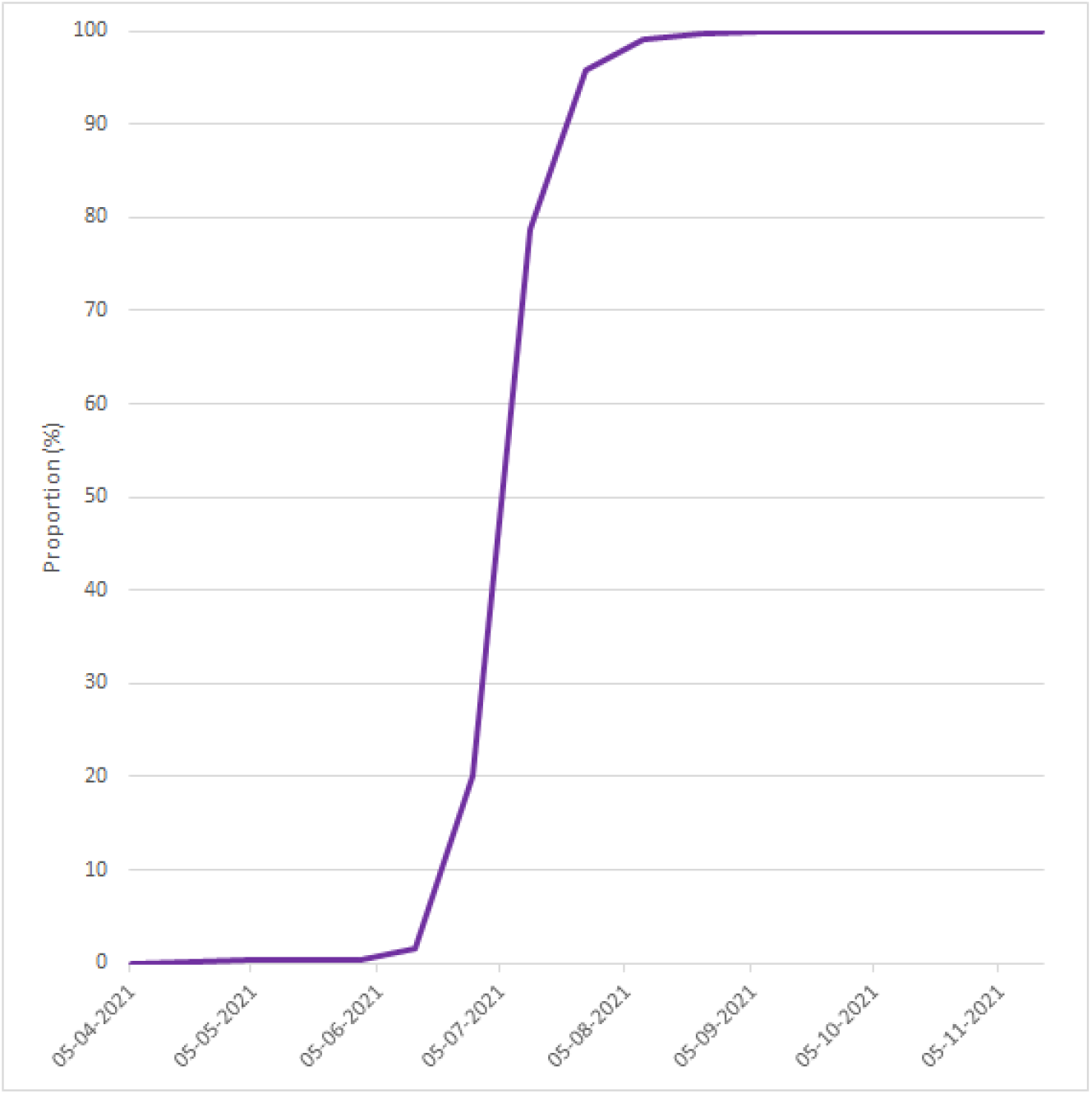
Share of Cases with Delta VOC Notes: This figure shows the proportion of sequenced positive samples that were identified as the Delta VOC in Denmark, April to November, 2021. Data source: Ritchie et al. (2020).

Between June and July the Delta VOC became the dominant variant in Denmark, reaching approximately 100% of all new cases (Figure 3).

In the study period, vaccinations were rolled out, going from below 40% fully vaccinated to more than 75% fully vaccinated individuals (Figure 4).

**Figure 4:**
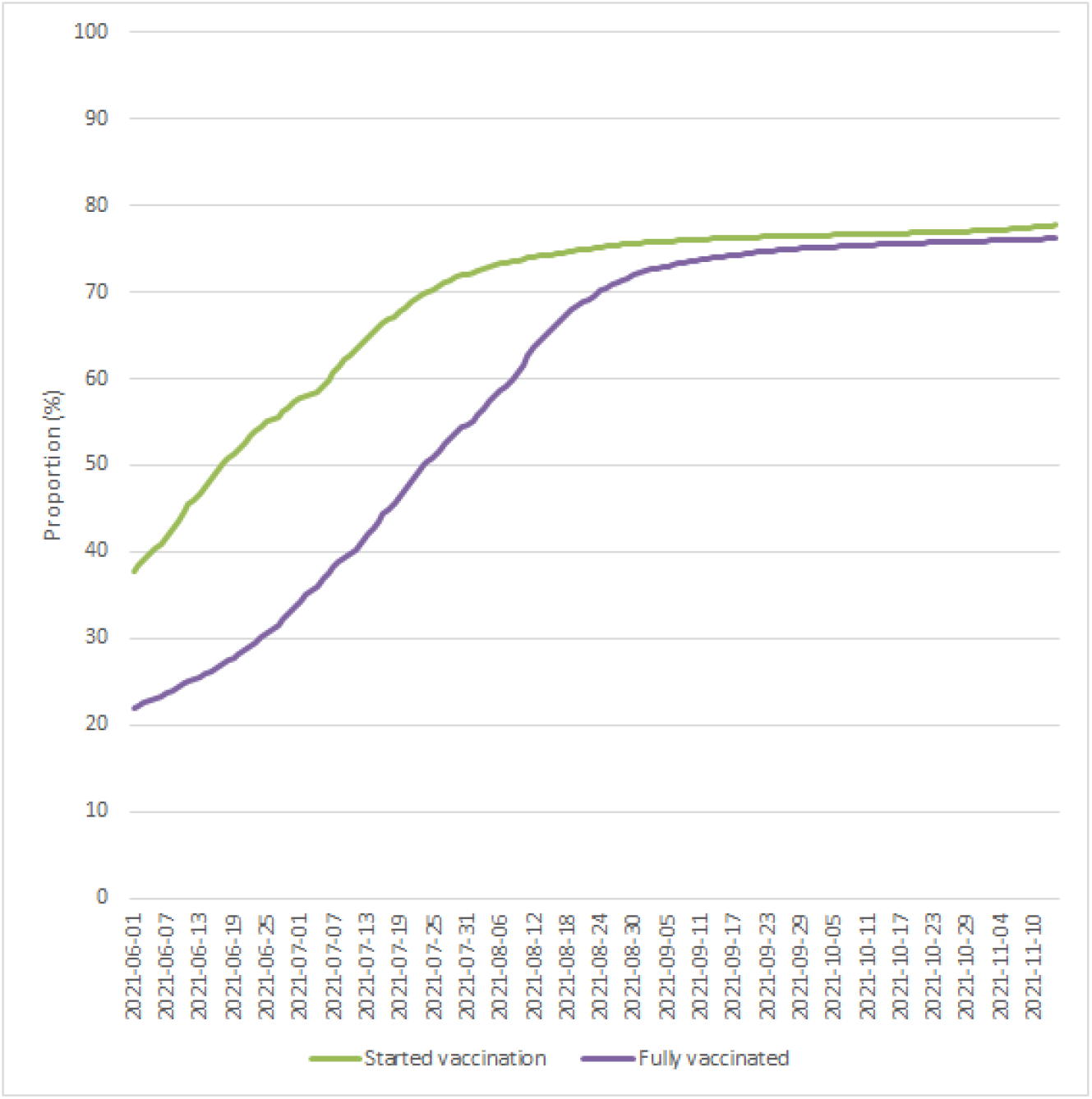
Proportion of population vaccinated Notes: This figure shows the proportion of the population partially and fully vaccinated, June to November 2021. Data source: SSI (2021b).

Denmark had a vaccination roll-out strategy with prioritization of vulnerable people. Nursery home residents were prioritized (group 01), followed by citizens above age 65 who had the need of personal help and care (group 02), then elderly people above age 85 (group 03). The next three groups (04-06) contained employees in healthcare and social work, high-risk patients, and relatives of high-risk patients. The remaining population was subsequently prioritized based on birth year (groups 07-17). Naturally, this leads to a correlation between calendar time and vaccination status conditional on age. Figure 5 shows the proportion of each group being fully vaccinated from January to November, 2021. We see that the compliance is generally high, and fast roll-out is seen in all groups. This roll-out strategy also meant that most of the vulnerable population was fully vaccinated, when the Delta VOC was first detected in Denmark. For example, more than 90% of individuals above age 65 (group 09) were fully vaccinated.

**Figure 5:**
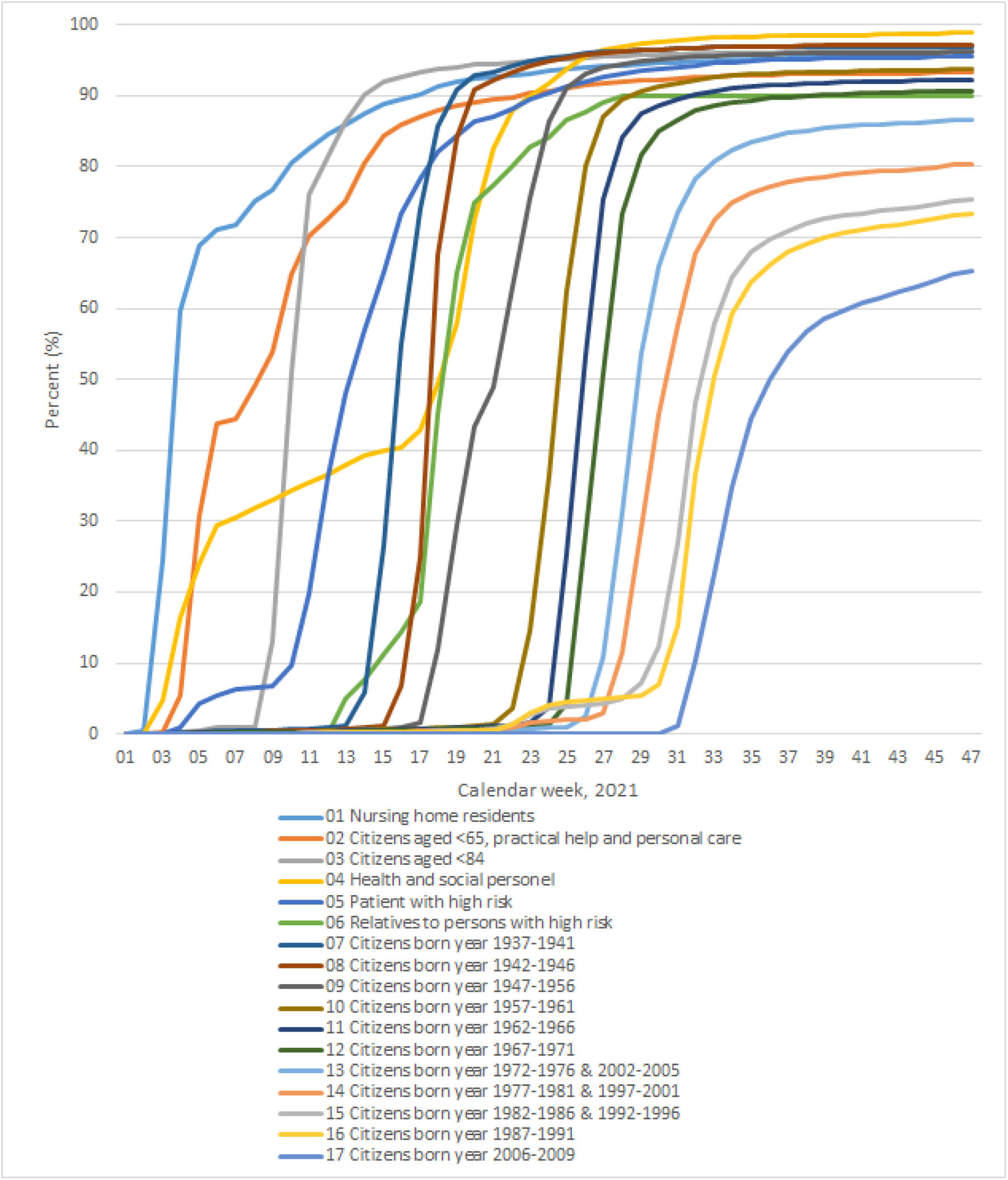
Vaccination roll-out Notes: This figure shows the proportion of the vaccination groups that are fully vaccinated by calendar week, January to November, 2021. Data source: SSI (2021b).

Finally, Denmark had relatively lenient restrictions during our study period. Figure 6 shows the government stringency index between June and November, 2021. The index is a composite measure of the strictness of policy responses and is based on nine response indicators including school closures, workplace closures, and travel bans. The index is calculated by the Oxford COVID-19 Government Response Tracker.

**Figure 6:**
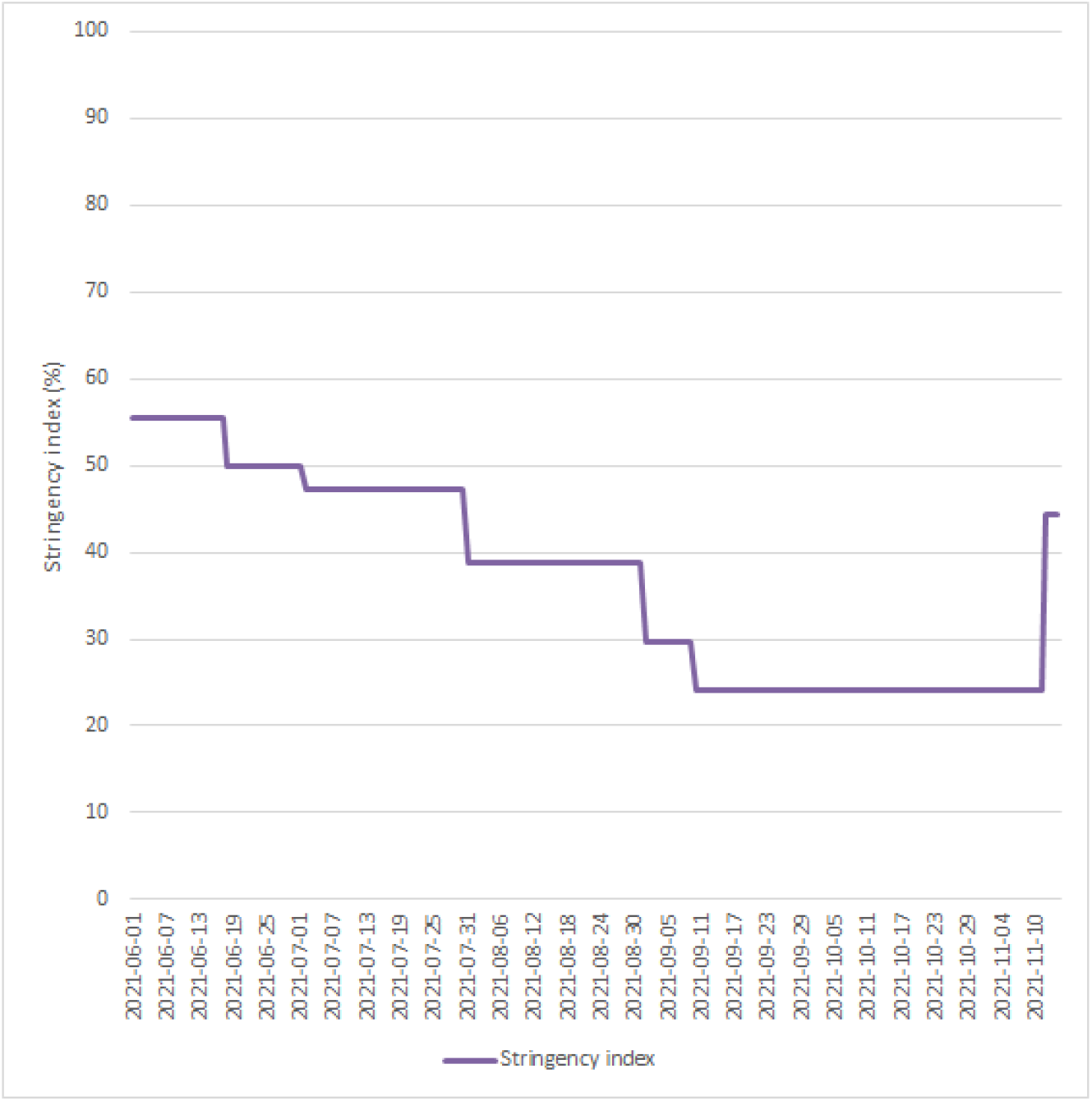
Government stringency index Notes: This figure shows the government stringency index June to November, 2021. The index is a composite measure of the strictness of policy responses and is based on nine response indicators including school closures, workplace closures, and travel bans. The index is calculated by the Oxford COVID-19 Government Response Tracker. Data source: Ritchie et al. (2020).

**Figure 7:**
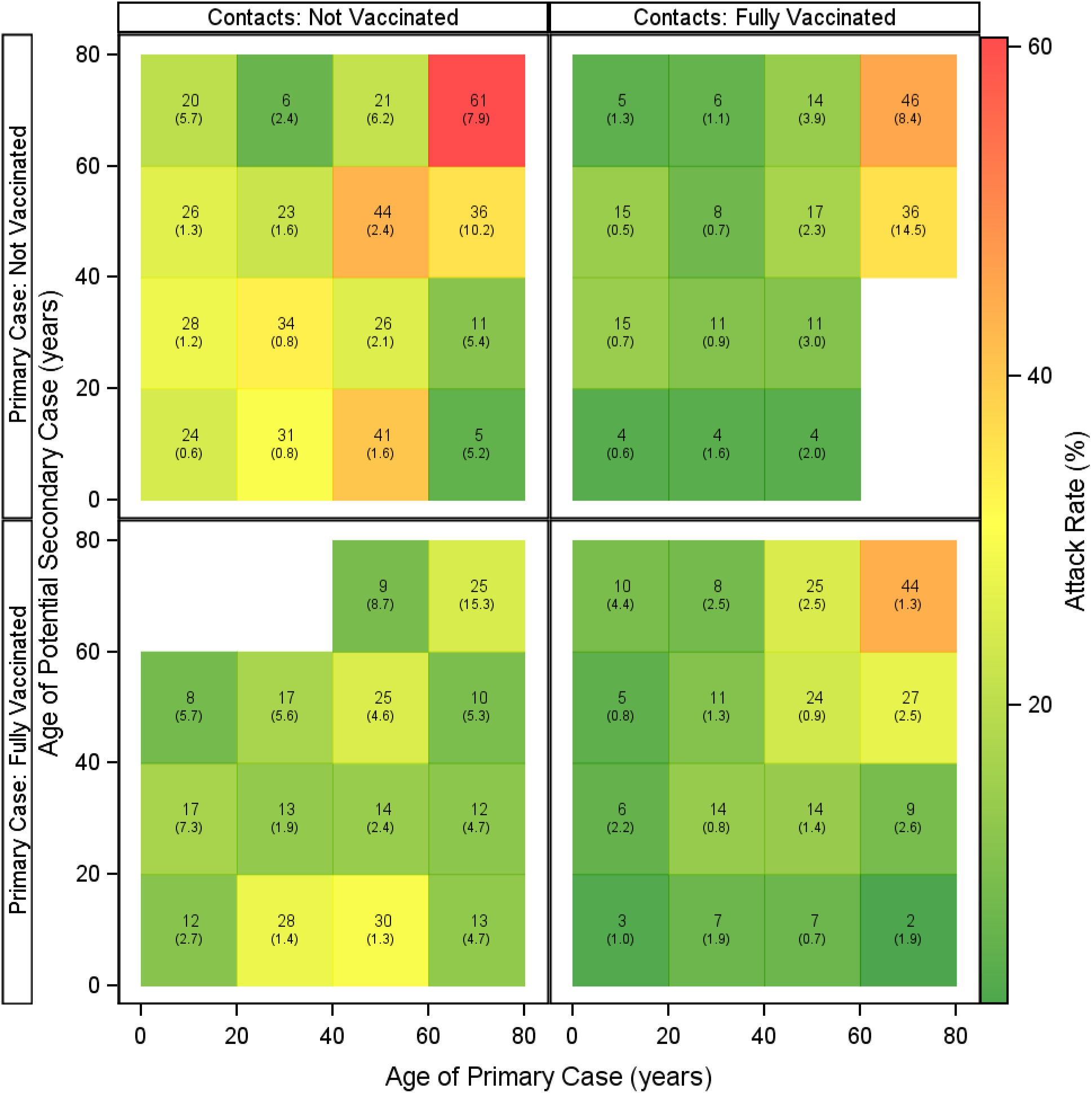
Secondary Attack Rate, age-by-age, stratified by vaccination status. Notes: This figure presents attack rates stratified by the age of the primary and potential secondary case, stratified by the vaccination status of the primary and potential secondary case. Numbers show the estimated secondary attack rates with 95% confidence intervals clustered on the household level in parentheses. Table 4 provides number of observations used to estimate the SAR for each combination.

##### 6.1 Danish testing program

The Danish SARS-CoV-2 testing program is based on easily accessible testing that is free of charge. The use of testing has been broadly accepted by the public and by December 2021, more than 15% of the Danish population were RT-PCR tested each week and a similar percentage were antigen tested. There are three tracks in the testing program: The healthcare track; the community track; and the rapid test track. Both the health-care track and the community track are based on RT-PCR detection of SARS-CoV-2 in oropharyngeal swabs, whereas the rapid test track is based on antigen detection. The RT-PCR results are usually reported within 24 hours. Individuals that test positive in the rapid test track are recommended to take a follow up RT-PCR test. All positive test results are registered in the Danish national microbiology database (MiBa), which thus provides a comprehensive overview of the pandemic on a national level.

##### 6.2 Genomic data

All RT-PCR-positive samples in the study period were selected for whole-genome sequencing by The Danish COVID-19 Genome Consortium (DCGC, 2021) if they showed a Ct value below 35 (Lyngse et al., 2021a). Whole-genome sequencing of the positive samples was performed on short read Illumina technology (Illumina, San Diego, California, United States) with the Illumina COVIDSeq Test kit (Illumina, San Diego, California, United States). The library preparation was performed as described by the manufacturer with a few modifications: COVIDSeq primer pool 2 was spiked with 1x amplicon 64 primer pair and 0.2x amplicon 70 and 74 primer pairs from the ARTIC v3 amplicon sequencing panel (ARTIC, 2022). Furthermore, in the cDNA amplification, the denaturation was adjusted to 95°C for 15 seconds and annealing at 63°C for 5 min. Samples were pooled and sequenced in batches for 384 on the Illumina NextSeq 500/550 using the mid-output kit v2.5 (150 cycles) kit, or 2×384 on the Illumina NovaSeq 6000 using the SP reagent kit (Illumina, San Diego, California, United States). Consensus sequences were called using “IVAR consensus”, with an in-house implementation of IVAR (version 1.3.1). The resulting consensus sequences were considered for variant calling when containing <3,000 ambiguous sites including N’s. Variants were called using Pangolin (version 3.1.16) with PangoLEARN assignment algorithm on the consensus sequences (O’Toole et al., 2021).

#### 7 Additional Analyses

##### 7.1 Age-by-age transmission

To investigate the age-related transmission patterns, we split the data into 20-year age groups of both the primary cases and potential secondary cases and estimated the secondary attack rate between all combinations of age groups, stratified by vaccination status of both the primary and potential secondary case (Figure 7). Generally, the SAR was highest, when both the primary and potential secondary case were unvaccinated (top-left panel). Furthermore, the SAR was generally higher when the primary case was vaccinated and the secondary case was not vaccinated (bottom left panel), than when the opposite was true (top right panel), showing that vaccination overall has a higher VES than VET, corroborating the findings in Table 2.

Next, we estimated the pooled VES, pooled VET, and VEC for each combination of age group (Figure 8). Generally, there was a positive VE across all age group combinations, implying that the SAR is reduced by vaccination of both the susceptible and infectious individual. Also, there was generally a decreasing VE with age of both the primary and potential secondary case, which could be related to waning immunity.

**Figure 8:**
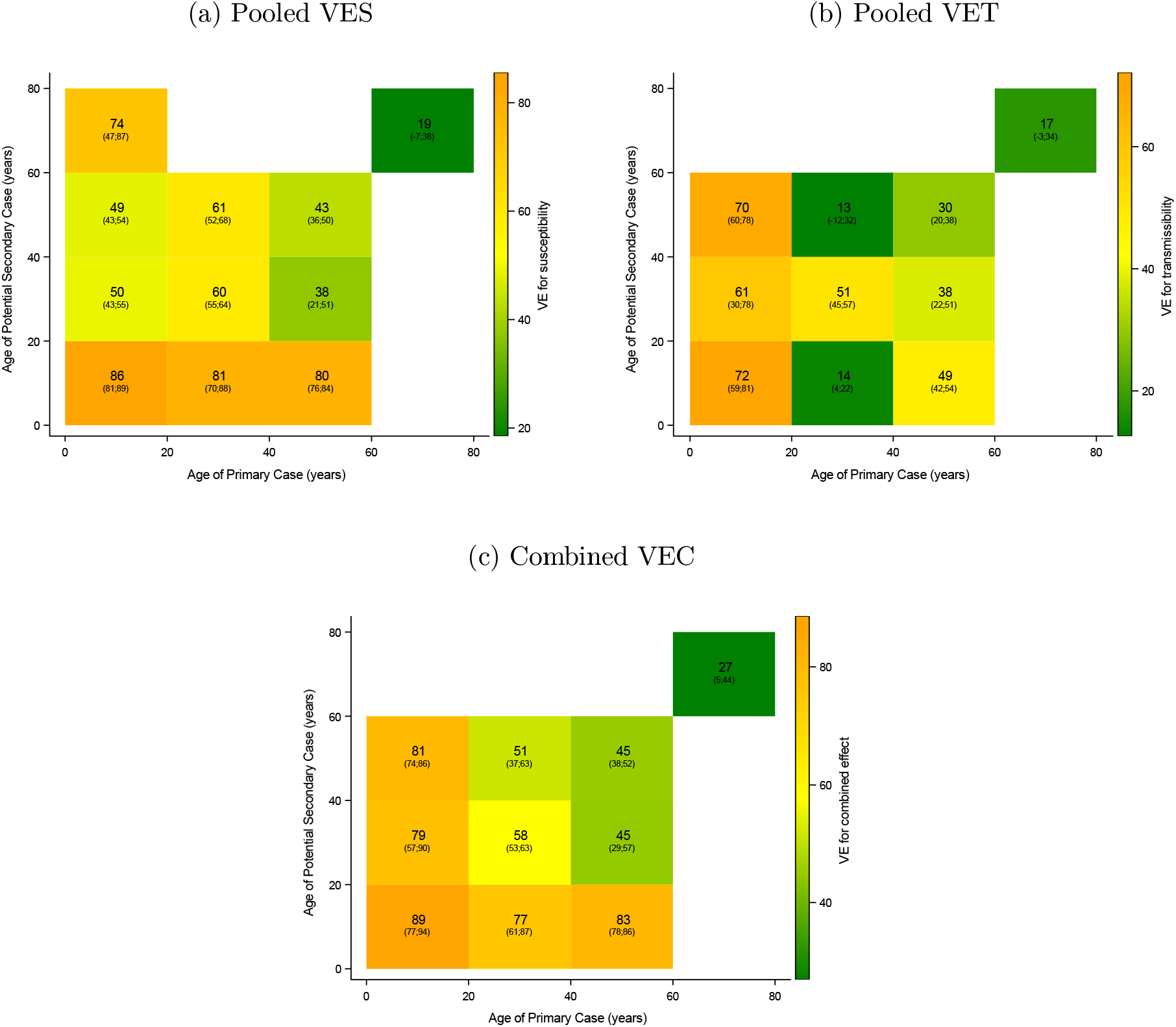
Crude VE estimates, stratified by age of the primary case and potential secondary case. Notes: This figure shows crude VE estimates stratified by age of the primary and potential secondary case. 95% confidence intervals clustered on the household level in parentheses. Panel a shows the pooled VES. Panel b shows the pooled VET. Panel c shows the combined effect, VEC.

##### 7.2 Intra-household correlation of lineages

We find an overall intra-household correlation of SARS-CoV-2 lineages of 88% (95%-CI: 87-89) (Table 5), i.e., 88% of all positive secondary cases had the same lineage as the primary case within the same household (conditional on the secondary test sample having a successfully sequenced genome). This intra-household correlation is lower compared to the one found in Lyngse et al. (2021a), at 96-99%. This could be because the society is more open, so there is a higher risk of community infection relative to household infection. However, it could also be due to uncertainty in the classification of subtype lineages of the Delta VOC, which is changing over time. In the present study, we used PANGO Lineage classifications from 2021-11-10.

In the present study, there is potential bias in the intra-household correlation of lineages across vaccinated and unvaccinated individuals, which could invalidate our comparisons of the relative risks. Table 5 presents the intra-household correlation of lineages across all combinations of vaccinated and unvaccinated primary and secondary cases. We found no statistically significant difference across any of the combinations, so there is no evidence for differential bias across our groups of comparison.

##### 7.3 Correlation of vaccination status within households

Vaccination status among household members are likely correlated due to several reasons. First, individuals are probably likely to live with other individuals with the same beliefs. Thus individuals that are pro being vaccinated are more likely to live together with other individuals that also are pro being vaccinated—and vice versa. Second, individuals are likely to live with a partner around their own age. As vaccination eligibility is correlated with age, individuals are likely to be eligible for vaccination around the same time. Third, there may be a fixed cost of being vaccinated, e.g., transportation time to the vaccination place. Thus, households may pool their time of vaccination on the same day to minimize these costs.

Figure 9 provides estimates of the correlation of the vaccination status between primary cases and potential secondary cases, stratified by five year age groups. It clearly shows some correlation among household members. Among young children (<12 years) there is perfect correlation, as they were not eligible for being vaccinated. We found a correlation of 0.63 within the full sample and a correlation of 0.72, when restricting the sample to individuals above age 12.

**Figure 9:**
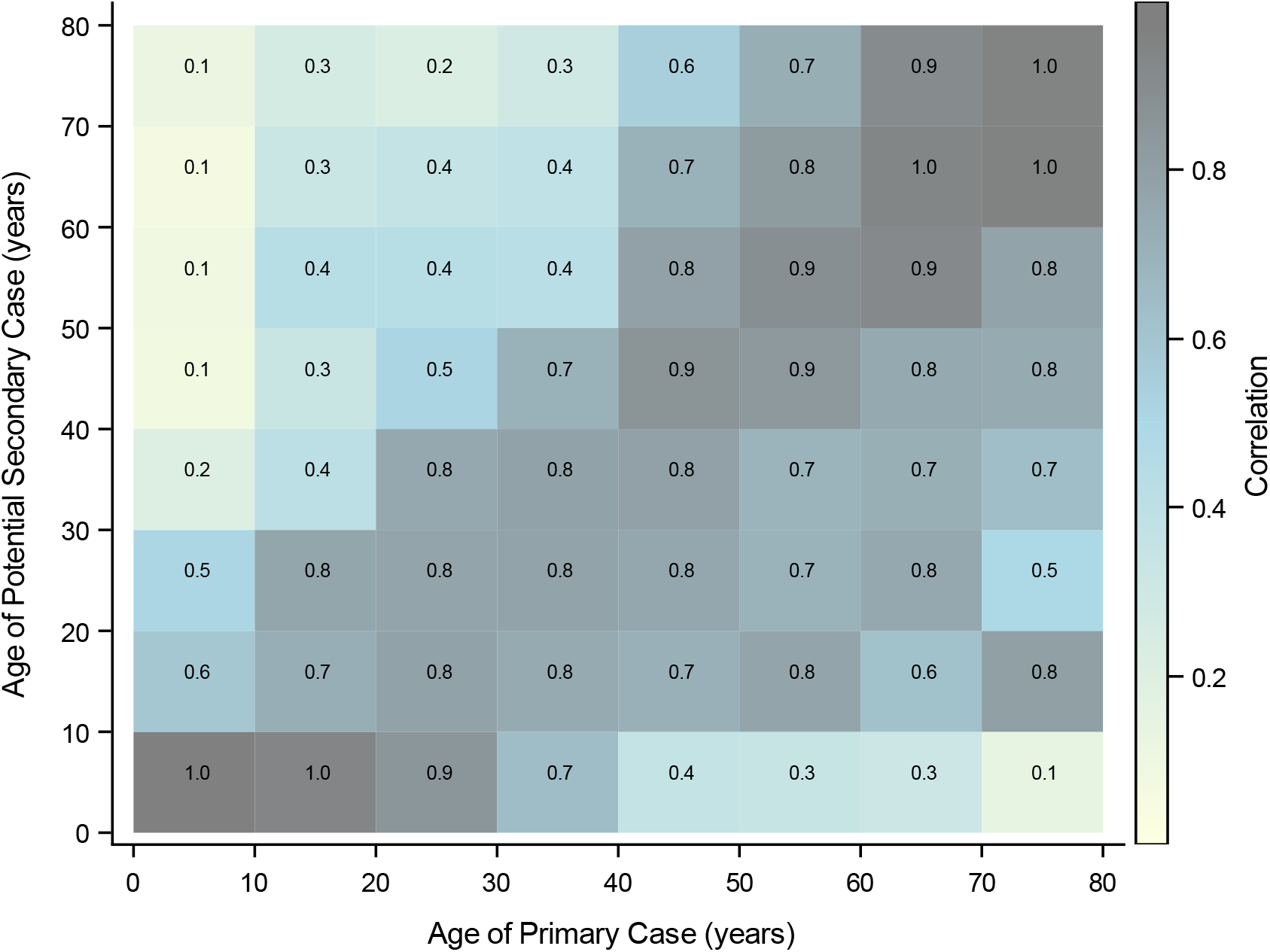
Correlation of vaccination status within households Notes: This figures provides estimates of the correlation of the vaccination status between primary cases and potential secondary cases, stratified by ten-year age groups.

**Figure 10:**
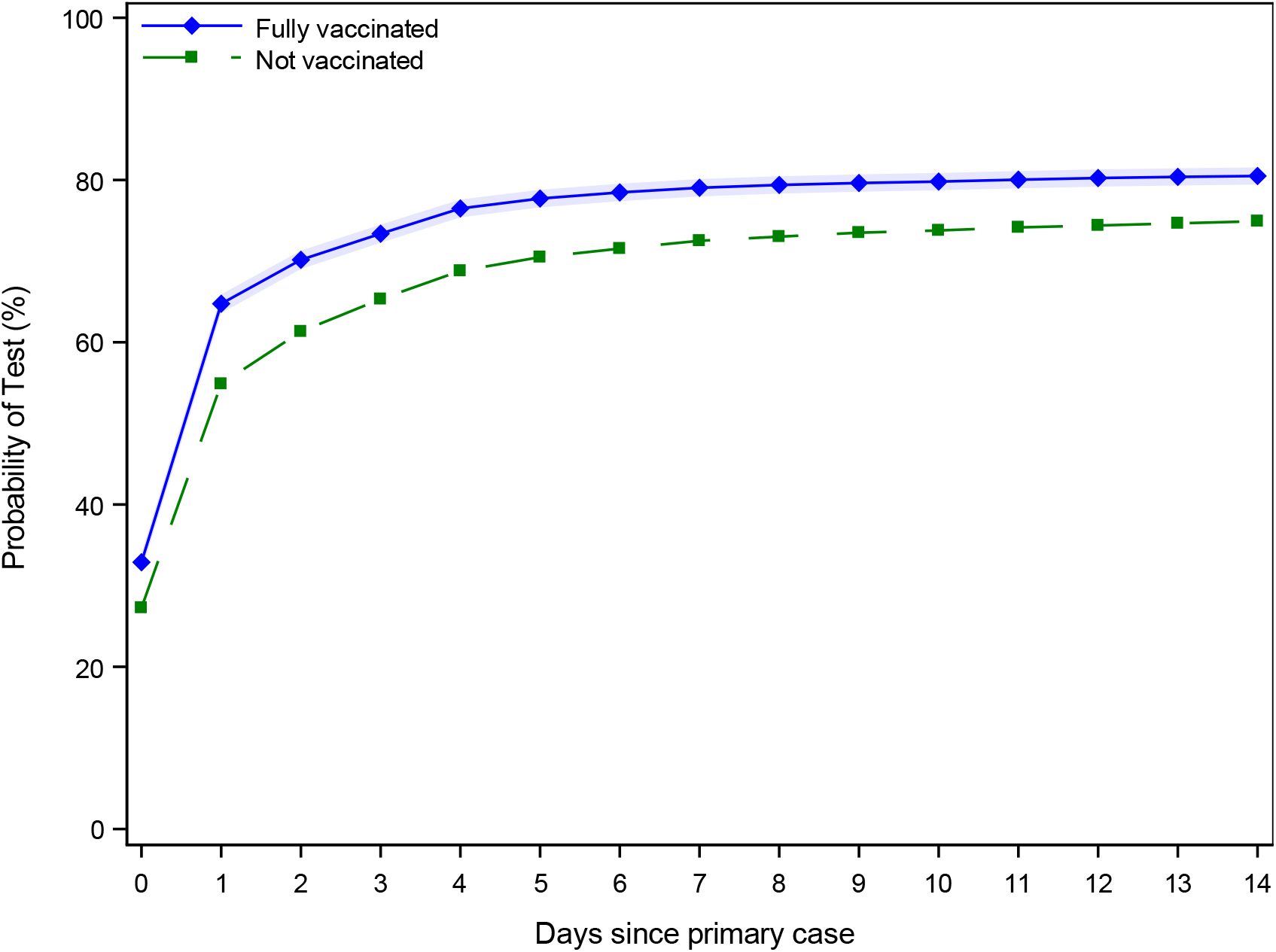
Probability of being tested after household exposure Notes: This figure provides estimates of the difference in the probability that a fully vaccinated potential secondary cases have been tested after identification of a primary case compared with an unvaccinated potential secondary case. It shows that the potential secondary cases are exhibiting the same trend in testing since vaccination, unconditional of the vaccination status. Vaccinated potential secondary cases have a significantly higher rate of being tested after diagnosis of a primary case in the same household. Shaded areas are 95%-confidence intervals clustered on the household level.

##### 7.4 Testing probabilities across vaccinated and unvaccinated individuals

When estimating the probability that an individual tests positive, it is conditional on the individual actually being tested. Selection bias is a potential concern, if the vaccination status of a potential secondary case within the household is correlated with the probability of being tested after the identification of the primary case. Overall, we find that potential secondary cases that are fully vaccinated are 7 percentage points (on a basis of 75%) more likely to be tested 1-14 days after exposure, compared to unvaccinated individuals (Table 10). Furthermore, potential secondary cases being exposed to primary cases that are fully vaccinated are also more likely to be tested. This suggests that there is a correlation between the household vaccination status and the compliance with being tested after exposure to a close contact.

##### 7.5 Ct values for vaccinated vs unvaccinated cases

One concern in investigating the transmissibility among vaccinated and unvaccinated cases is that the viral load may differ across the two groups. Indeed the literature has shown that the viral load can be reduced for vaccinated individuals with breakthrough infections compared to unvaccinated individuals (Levine-Tiefenbrun et al., 2021). We found that samples from vaccinated primary cases had a lower viral load distribution (higher Ct values) compared to samples from unvaccinated primary cases (Figure 11).

**Figure 11:**
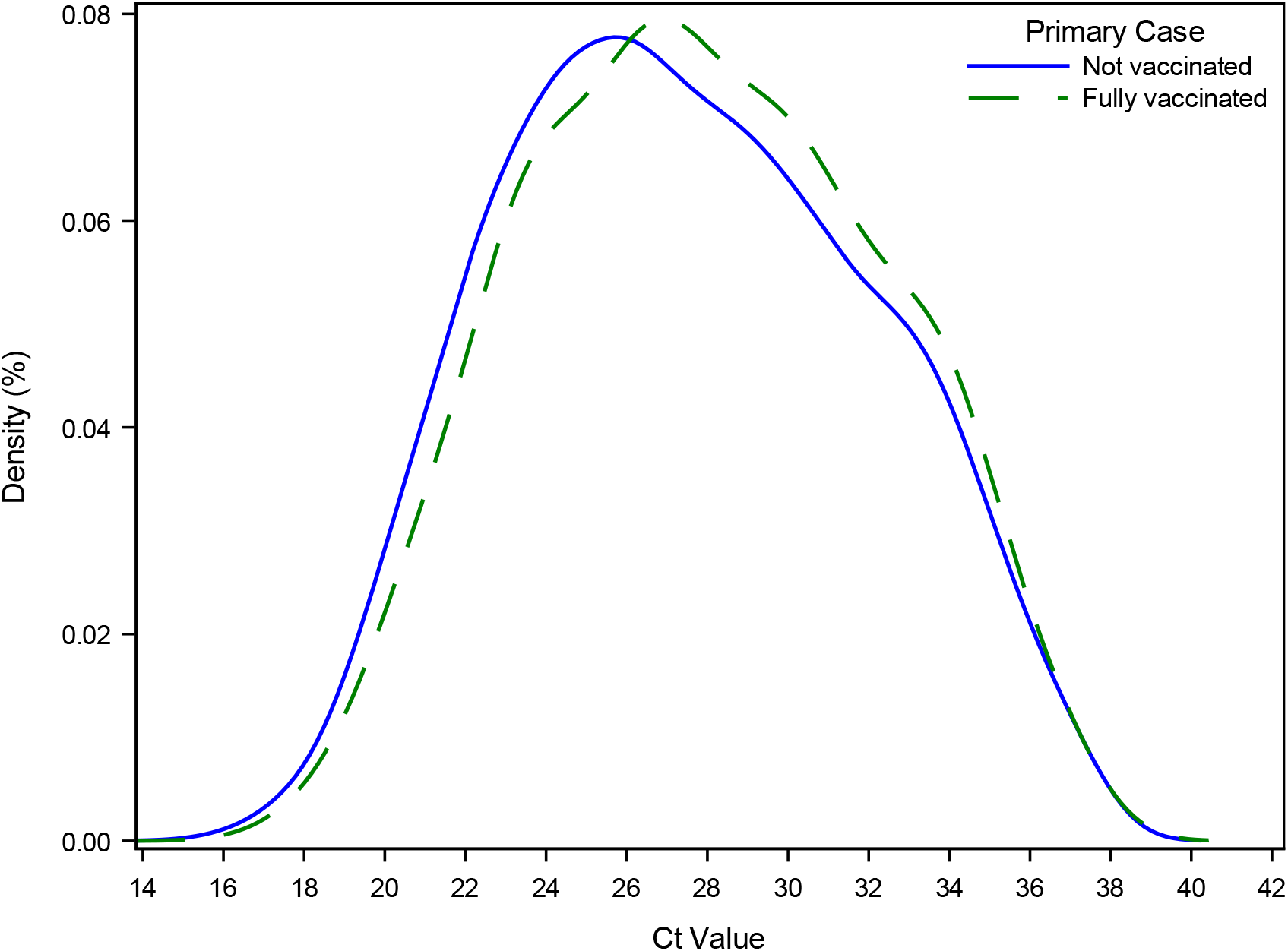
Ct values across vaccinated and unvaccinated primary cases Notes: This figure shows the density of the Ct values of primary cases stratified by the vaccination status. Samples from vaccinated cases had a lower viral load (higher Ct values) distribution compared to samples from unvaccinated primary cases.

However, this result may be biased if, for example, vaccinated cases with a breakthrough infection are being diagnosed later in their infection compared to unvaccinated cases. This could for example occur if vaccinated individuals are tested less regularly than unvaccinated individuals, if they are asymptomatic or do not develop as severe symptoms, or if they believe they are less likely to be infected because of their vaccination status. We investigated this potential selection bias by comparing secondary cases that were fully vaccinated with unvaccinated secondary cases that tested positive on the same day after exposure of the primary case (Figure 1 and Table 6). We found that vaccinated secondary cases had a lower viral load—a 1.6 point higher Ct value, translating into a third of a standard deviation.

##### 7.6 Robustness of VE estimates

This section addresses the robustness of the VE estimates presented in Table 2. Table 7 provides VE estimates, conditional on the potential secondary case having a test result. Thus, we do not assume that all untested contacts are negative. Table 8 provides VE estimates, including Ct values of the primary case as a proxy for viral load. Table 9 provides VE estimates, including Ct values of the primary case as a proxy for viral load, conditional on the potential secondary case having a test result. Table 10 provides VE estimates stratified by time since vaccination, in order to take waning immunity into account. Table 11 provides VE estimates stratified by time since vaccination, in order to take waning immunity into account, conditional on the potential secondary case having a test result.

**Table 6:** Comparison of Ct values between vaccinated and unvaccinated secondary cases

|  | I | II | III | IV |
| --- | --- | --- | --- | --- |
| Fully vaccinated | 0.29<br>(0.8882) | 1.61<br>(0.8655) | 1.63<br>(0.1843) | 1.30<br>(0.2458) |
| Unvaccinated, mean | 28.95 | 28.95 | 28.95 | 28.95 |
| Unvaccinated, STD | 4.74 | 4.74 | 4.74 | 4.74 |
| Estimate / STD | 0.06 | 0.34 | 0.34 | 0.27 |
| Days since primary case FE | YES | YES | YES | YES |
| Age FE | NO | YES | YES | YES |
| Age, primary case, FE | NO | NO | YES | YES |
| Fully vaccinated index | NO | NO | YES | YES |
| Ct value index | NO | NO | NO | YES |
| N Observations | 6,938 | 6,938 | 6,938 | 4,091 |
| N households | 5,088 | 5,088 | 5,088 | 2,963 |

Notes: This table provides regression estimates for the Ct value for secondary cases, comparing unvaccinated cases with vaccinated cases, controlling for the day of testing since exposure to the primary case and 5 year age fixed effects. FE = Fixed Effects.

**Table 7:** Vaccine effectiveness, conditional on potential secondary case having a test result

|  | Susceptibility |  |  | Transmissibility |  |  | Combined |
| --- | --- | --- | --- | --- | --- | --- | --- |
| Primary cases vaccinated | Pool | Not | Fully |  |  |  |  |
| Potential secondary cases vaccinated |  |  |  | Pool | Not | Fully |  |
| Estimator | $VES_{N,V/N,N}$ | $VES_{V,V/V,N}$ | $VES_{.,V/.,N}$ | $VET_{V,N/N,N}$ | $VET_{V,V/N,V}$ | $VET_{V.,/N.,}$ | $VEC_{V,V/N,N}$ |
| VE (%) | 65 | 61 | 61 | 51 | 36 | 28 | 72 |
| (95%-CI) | (64;67) | (59;64) | (56;65) | (48;53) | (32;40) | (20;35) | (70;73) |
| Age FE | YES | YES | YES | YES | YES | YES | YES |
| Week FE | YES | YES | YES | YES | YES | YES | YES |
| Female | YES | YES | YES | YES | YES | YES | YES |
| Household size | YES | YES | YES | YES | YES | YES | YES |
| Female, primary case | YES | YES | YES | YES | YES | YES | YES |
| Age, primary case, FE | YES | YES | YES | YES | YES | YES | YES |
| Ct value, primary case, FE | NO | NO | NO | NO | NO | NO | NO |
| Conditional on test | YES | YES | YES | YES | YES | YES | YES |
| N Observations | 42,170 | 29,472 | 12,698 | 42,170 | 20,592 | 21,578 | 26,310 |
| N Households | 21,086 | 13,698 | 7,388 | 21,086 | 12,744 | 14,201 | 17,220 |

Notes: This table provides VE estimates from the same model as Table 2, conditional on the potential secondary case having a test result. The estimates of vaccine effectiveness against transmissibility (VET) is given as a pooled estimate and stratified by the vaccination status of the potential secondary cases within the household. The combined vaccine effectiveness (VEC) is defined as both the primary and potential secondary case being vaccinated relative to them both being unvaccinated. Vaccine effectiveness is estimated as one minus the relative risk of vaccinated individuals relative to unvaccinated individuals. Note that the VE estimates across columns are not directly comparable as they are estimated on stratified samples. 95% confidence intervals clustered on the household level in parentheses. FE = included as fixed effects in the model.

**Table 8:** Vaccine effectiveness, controlling for primary case Ct values

|  | Susceptibility |  |  | Transmissibility |  |  | Combined |
| --- | --- | --- | --- | --- | --- | --- | --- |
| Primary cases vaccinated | Pool | Not | Fully |  |  |  |  |
| Potential secondary cases vaccinated |  |  |  | Pool | Not | Fully |  |
| Estimator | $VES_{N,V/N,N}$ | $VES_{V,V/V,N}$ | $VES_{.,V/.,N}$ | $VET_{V,N/N,N}$ | $VET_{V,V/N,V}$ | $VET_{V.,/N.,}$ | $VEC_{V,V/N,N}$ |
| VE (%) | 61 | 62 | 47 | 40 | 30 | 4 | 66 |
| (95%-CI) | (58;63) | (58;64) | (38;54) | (35;45) | (23;36) | (-10;17) | (62;69) |
| Age FE | YES | YES | YES | YES | YES | YES | YES |
| Week FE | YES | YES | YES | YES | YES | YES | YES |
| Female | YES | YES | YES | YES | YES | YES | YES |
| Household size | YES | YES | YES | YES | YES | YES | YES |
| Female, primary case | YES | YES | YES | YES | YES | YES | YES |
| Age, primary case, FE | YES | YES | YES | YES | YES | YES | YES |
| Ct value, primary case, FE | YES | YES | YES | YES | YES | YES | YES |
| Conditional on test | NO | NO | NO | NO | NO | NO | NO |
| N Observations | 28,364 | 20,148 | 7,917 | 28,364 | 14,236 | 14,128 | 17,564 |
| N Households | 13,056 | 8,577 | 4,311 | 13,056 | 8,162 | 8,898 | 10,627 |

Notes: This table provides VE estimates from the same model as Table 2, controlling for the sample Ct value of the primary case. The estimates of vaccine effectiveness against transmissibility (VET) is given as a pooled estimate and stratified by the vaccination status of the potential secondary cases within the household. The combined vaccine effectiveness (VEC) is defined as both the primary and potential secondary case being vaccinated relative to them both being unvaccinated. Vaccine effectiveness is estimated as one minus the relative risk of vaccinated individuals relative to unvaccinated individuals. Note that the VE estimates across columns are not directly comparable as they are estimated on stratified samples. 95% confidence intervals clustered on the household level in parentheses. FE = included as fixed effects in the model. VES for primary cases fully vaccinated only includes primary cases with a sample Ct value >20.

**Table 9:**
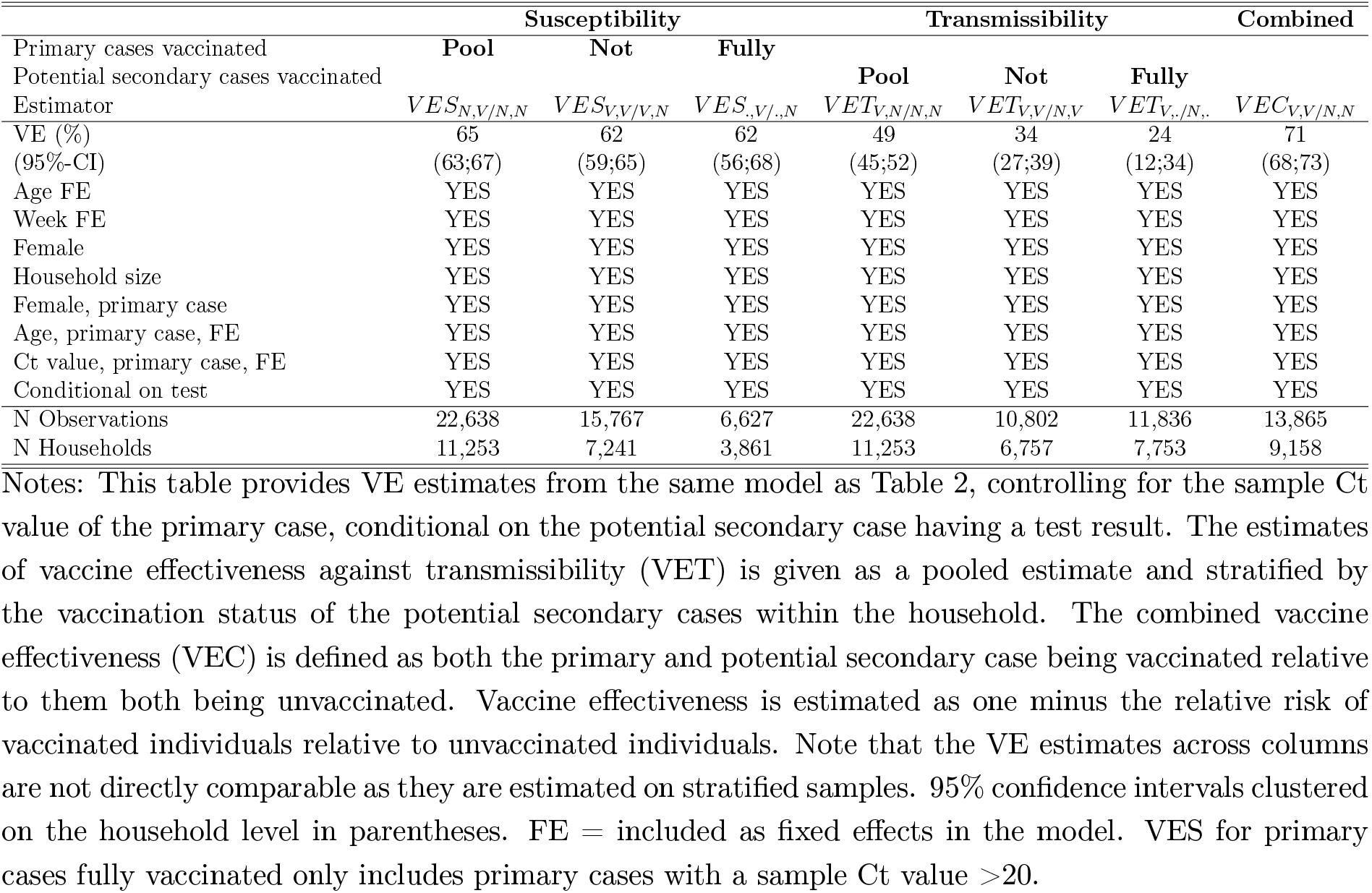
Vaccine effectiveness, controlling for primary case Ct values, conditional on potential secondary case having a test result

**Table 10:**
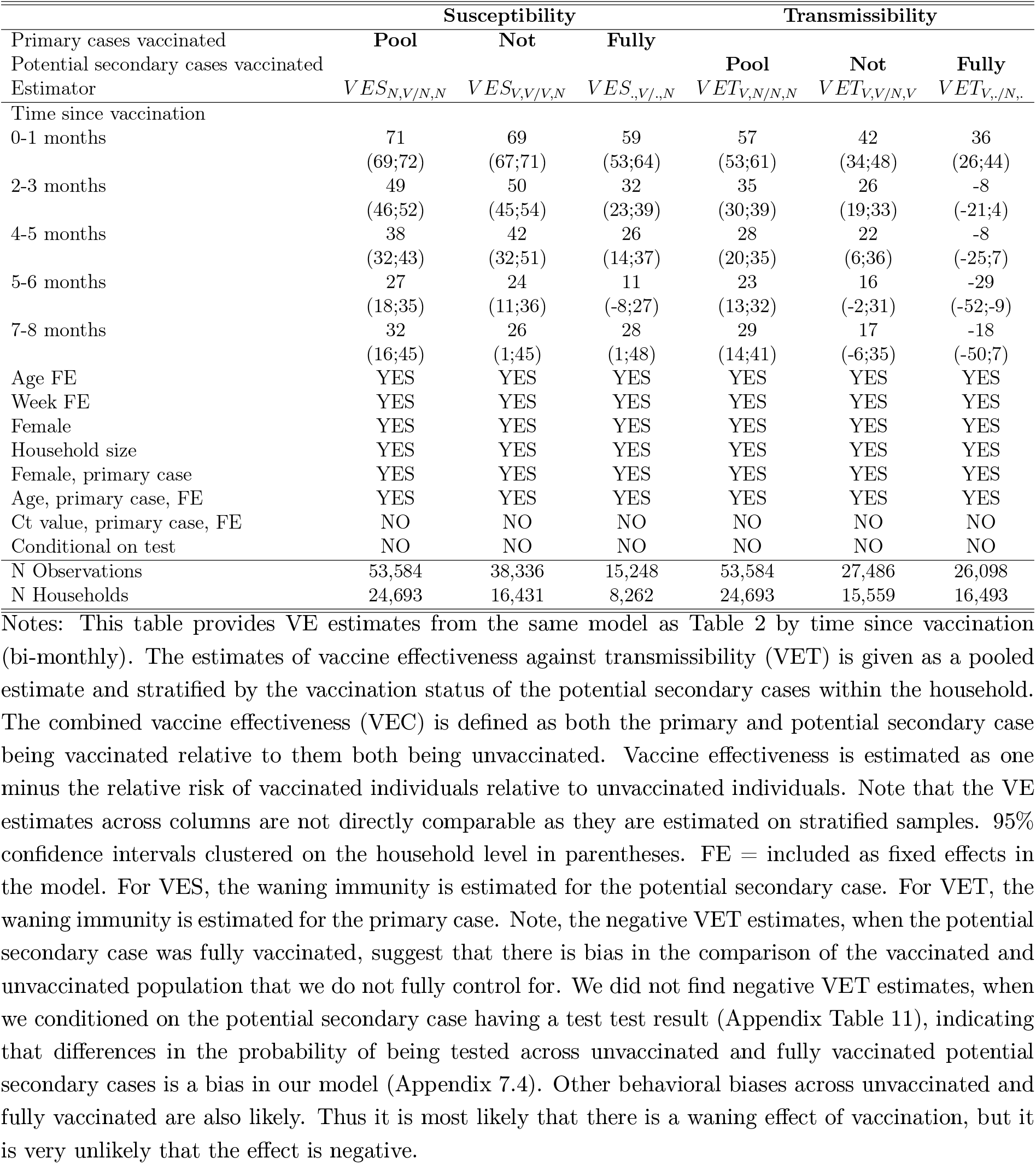
Vaccine effectiveness by time since vaccination

**Table 11:**
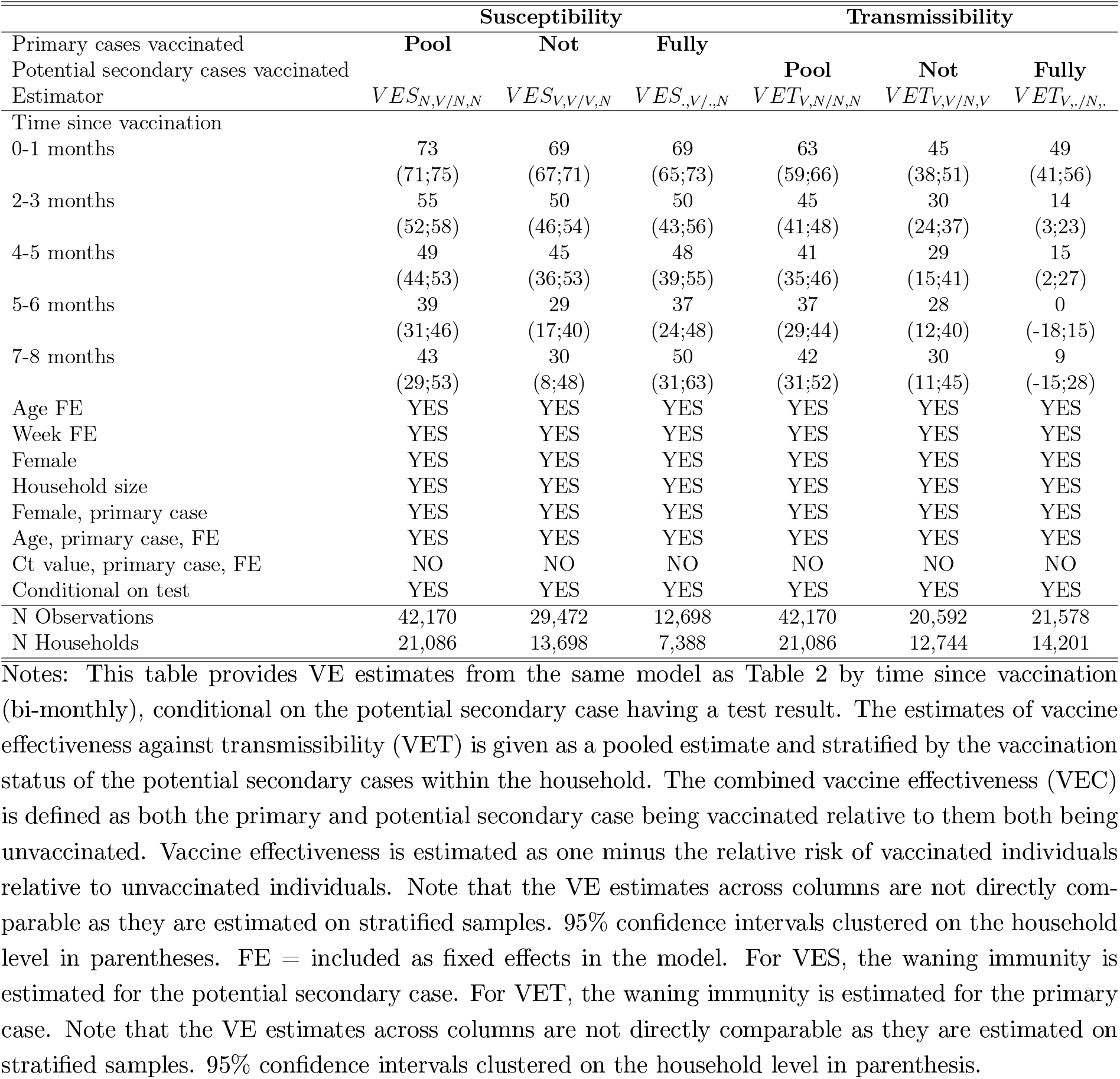
Vaccine effectiveness by time since vaccination, conditional on potential secondary case having a test result

#### 8 Statistical Appendix

This section provides more details of the statistical methods used to generate the results presented in the main manuscript.

We defined the secondary attack rate (SAR) as the proportion of potential secondary cases that were infected within each household 1-14 days after exposure to a primary case. We only included individuals that were either fully vaccinated (V) or not vaccinated (N), thus excluding individuals that were partially vaccinated.

To compare attack rates across different vaccination status of primary cases and potential secondary cases, we estimated the relative risk (RR). The vaccine effectiveness (VE) is given by one minus the relative risk (1-RR) of the SAR of vaccinated individuals compared to the SAR of the unvaccinated individuals. We stratified our analyses in order to separate the effect of susceptibility (VES), the effect of transmissibility (VET), and the combined effect (VEC). In particular, we use the following 7 equations, with underlying estimates for SAR produced using generalized linear models that control for age, sex, household size, and calendar week, with standard errors clustered on the household level.

Let 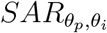 denote the SAR, where *θ* is the vaccination status *θ* ∈ {*V, N*} (*Fully Vaccinated* or *Not vaccinated*) of primary cases (*p*) and potential secondary cases (*i*). Let “.” denote the pooled sample of both vaccination statuses. Thus, the pooled SAR is denoted as *SAR*_.,._

##### Vaccine effectiveness of susceptibility (*V ES*)

First, we compared the SAR across potential secondary cases that were fully vaccinated and potential secondary cases that were not vaccinated, unconditional of the vaccination status of the primary case.

The pooled vaccine effectiveness against susceptibility (*V ES*) is given by:

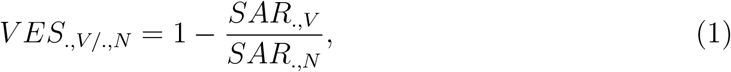

i.e., the SAR of vaccinated potential secondary cases relative to the SAR of unvaccinated potential secondary cases.

Next, we hold the vaccination status of the primary case fixed, and compare the SAR across potential secondary cases that are fully vaccinated and not vaccinated.

Vaccine effectiveness of susceptibility among unvaccinated primary cases is given by:

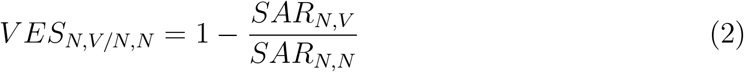

Vaccine effectiveness of susceptibility among fully vaccinated primary cases is given by:

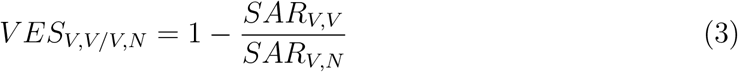

##### Vaccine effectiveness of transmissibility (*V ET*)

First, we compared the SAR across primary cases that were fully vaccinated and not vaccinated, unconditional of the vaccination status of the potential secondary cases.

The pooled vaccine effectiveness of transmissibility (*V ET*) is given by:

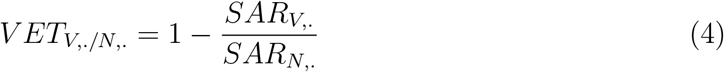

Next, we hold the vaccination status of the potential secondary cases fixed, and compare the SAR across primary cases that are fully vaccinated and not vaccinated.

Vaccine effectiveness of transmissibility among unvaccinated potential secondary cases is given by:

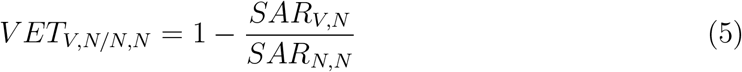

Vaccine effectiveness of transmissibility among fully vaccinated potential secondary cases is given by:

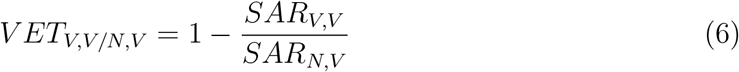

##### Combined effectiveness (*V EC*)

Finally, we compared the SAR, where both the primary and potential secondary cases were fully vaccinated, with the SAR, where both the primary and potential secondary cases were unvaccinated.

The combined vaccine effectiveness is given by:

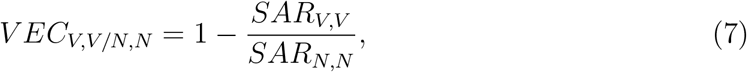

##### Estimation

We estimated the relative risks for VES, VET and VEC using the following generalized linear model, with a Poisson distribution response and log link function:

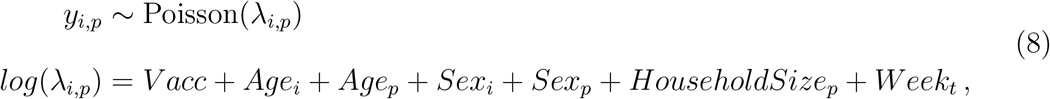

where *V acc* is a binary fixed effect representing one of the following explanatory variables for each model:

- For VES, *V acc* refers to vaccination status of the potential secondary case *i*.
- For VET, *V acc* refers to vaccination status of the primary case *p*.
- For VEC, *V acc* refers to vaccination status of both the primary and potential secondary case *p, i*.

*Age* denotes categorical fixed effects of age group in 10 year intervals, *Sex* is a binary fixed effect controlling for sex, *HouseholdSize* denotes categorical fixed effects of the number of household members, and *Week* is a categorical fixed effect of the calendar week *t*. Standard errors were clustered on the household level in order to control for within-household correlation in risk. Note that the Poisson distribution is used with binary outcome in order to facilitate the calculation of relative risks.

